# Multiple, but not isolated, yellow fever virus-associated orthoflavivirus immune histories drive antibody-dependent enhancement of Zika and dengue viruses

**DOI:** 10.64898/2026.05.22.26353817

**Authors:** Sebastian Gallon, Prince Baffour Tonto, Yifeng Ding, Guan-Hua Chen, Kailee Naito-Keoho, Carlos Brites, Eduardo Martins Netto, Wei-Kung Wang, Bobby Brooke Herrera

**Author notes:** **Corresponding Author**: BBH,; WKW.

## Abstract

Antibody-dependent enhancement (ADE) is a major concern across orthoflavivirus infections, yet how multiple viral exposures shape enhancement risk remains incompletely understood. Here, we integrated serosurveillance from Saúde, Brazil with functional immunologic analyses to define how yellow fever virus (YFV)-associated orthoflavivirus immune histories influence ADE phenotypes. Using serocomplex-specific anti-premembrane antibody profiling validated by microneutralization assays, plasma samples were stratified into YFV-only, YFV+DENV, and YFV+DENV+ZIKV exposure groups. In Fcγ receptor-bearing U937 cells, YFV-only plasma demonstrated minimal enhancement activity, whereas cumulative orthoflavivirus exposure generated broader ADE phenotypes across heterologous viruses. In *IFNAR1^−/−^* passive-transfer models, YFV-only plasma did not enhance ZIKV or DENV2 infection in vivo. In contrast, YFV+DENV plasma increased ZIKV viremia and accelerated mortality kinetics, while YFV+DENV+ZIKV plasma demonstrated concentration-dependent enhancement phenotypes. Collectively, these findings indicate that isolated YFV immunity does not predispose to ADE, whereas cumulative orthoflavivirus exposure generates antibody repertoires capable of producing concentration-dependent enhancement in vivo.

## Introduction

Orthoflaviviruses collectively account for millions of infections annually and represent a growing global public health concern. Dengue (DENV1-4), Zika (ZIKV), yellow fever (YFV), and West Nile (WNV) viruses, belonging to different serocomplexes, are responsible for widespread outbreaks, particularily across tropical and subtropical regions of the Global South ^1–5^. These viruses are transmitted primarily by two mosquito genera, including *Aedes*, which transmit DENV, ZIKV, and YFV, and *Culex*, which transmit WNV ^1–5^. Given the presence of mosquitoes in the tropics/subtropics and geographic overlap of these viruses, many populations in endemic regions experience exposures to multiple orthoflaviviruses over their lifetimes ^6,7^.

The immunological consequences of such sequential exposures are complex. Orthoflaviviruses share considerable antigenic similarity, particularly within the envelope (E) protein, which is the principle target of neutralizing antibodies ^8,9^. As a result, infection with one orthoflavivirus frequently induces antibodies that cross-react with related viruses ^10–12^. These cross-reactive responses can be either protective or pathogenic, depending on infection order, time interval, antibody titer, antibody avidity, and epitope specificity ^13–17^. Prior studies have demonstrated that sequential orthoflavivirus infections can leave an antigen imprint, biasing subsequent antibody responses toward conserved epitopes on the E protein ^8,18,19^. Such imprinting can favor the expansion of cross-reactive antibodies, including weakly neutralizing antibodies that mediate antibody-dependent enhancement (ADE) ^8,20^. ADE is particularly well characterized in the context of DENV, where secondary infection with a heterologous serotype is associated with increased risk of severe disease ^21,22^. In this setting, pre-existing non-neutralizing or sub-neutralizing antibodies form immune complexes with virus particles and facilitate entry into Fcγ receptor-bearing mononuclear cells, leading to increased viral replication and systemic viremia ^23,24^.

Mechanistically, ADE has been extensively characterized using both in vitro and in vivo systems. Human Fcγ receptor-expressing cell lines, such as U937 and K562, have become common platforms for evaluating enhancement activity and for assessing vaccine-induced antibody responses ^25,26^. These systems consistently demonstrate that cross-reactive antibodies that fail to fully neutralize virus can instead promote infection when present at sub-neutralizing concentrations ^27^. Complementary studies in murine passive transfer models have further established that heterologous anti-DENV or anti-ZIKV sera can enhance subsequent infection, resulting in increased viremia, inflammatory responses, and mortality ^23,24,28,29^. Studies in non-human primates have demonstrated ADE against DENV2 with passive transfer of human plasma and DENV monoclonal antibodies, but ADE of ZIKV has not been definitvely shown to have as large of a clinical impact in this model ^30–34^. Human cohort studies similarly support an infection-order-dependent and antibody-titer-dependent risk of ADE, particularly among individuals with heterologous sequential DENV infections or ZIKV-DENV exposures ^14,15,35^. Despite these advances, the functional consequences of layered orthoflavivirus exposure histories, particularly those shaped by YFV infection or vaccination, remain poorly defined at both the serological and mechanistic levels.

This gap is notable because YFV vaccination or natural exposure are widespread across endemic regions of Africa and South America. Many individuals encounter DENV, ZIKV, or other orthoflaviviruses in the context of pre-existing YFV immunity or followed by YFV exposure ^36–38^. Immunization campaigns have increasingly recognized that pre-existing orthoflavivirus immunity can influence vaccine performance, as both ADE risk and altered immunogenicity remain key concerns ^39–44^. For example, prior YFV-17D immunity has been shown to modulate antibody responses to subsequent tick-borne encephalitis virus (TBEV) vaccination ^41^. In that setting, pre-existing YFV immunity led to an original antigenic sin-like response characterized by reduced TBEV-specific neutralization and preferential expansion of broadly cross-reactive antibodies ^41^. Additionally, priming with YFV before an inactivated ZIKV vaccination led to broader binding recognition to an array of orthoflavirus antigens ^45^. Although cross-reactive antibody depletion did not compromise neutralization against the homologous viruses in that study, the potential of these antibodies to mediate ADE against other clinically relevant orthoflaviviruses was not assessed.

Understanding how multiple orthoflavivirus exposures shape functional antibody landscapes is therefore critical for both pathogenesis and vaccine design, yet this has been difficult to study in endemic settings where extensive serological cross-reactivity obscures prior infection histories. We leveraged our serocomplex-specific anti-pre-membrane (prM) assay to conduct a serosurveillance study of 300 participants in Saúde, a small town in the Northeastern state of Bahia, Brazil, to reconstruct immune histories of orthoflavivirus exposure including DENV, ZIKV, YFV, and/or WNV and integrated the information with microneutralization, in vitro ADE assays, and in vivo passive-transfer studies to define how cumulative orthoflavivirus exposures reshape the balance between neutralizing and enhancing antibody activities in the context of YFV immunity. We hypothesized that distinct orthoflavivirus exposure profiles will generate functionally different antibody repertoires with predictable neutralization and enhancement phenotypes. In addition to samples from Brazil based on our serosruveillance study, we included samples from individuals with well-documented YFV-17D vaccination and stratified them into YFV-only, YFV+DENV, and YFV+DENV+ZIKV groups and evaluated their functional consequences across multiple orthoflaviviruses.

## Results

### Serosruveillance of orthoflavivirus exposure in Saúde, Brazil

To define prior orthoflavivirus exposures within the study population, we performed Western blot (WB) analysis using a previously validated anti-prM antibody profiling approach capable of distinguishing exposure to the four major orthoflavivirus serocomplexes represented in this study (DENV, ZIKV, WNV, and YFV), with sensitivities and specificities ranging from 88.9-91.7% and 92.5-99.2%, respectively ^46^. Participants were classified as having prior DENV infection, ZIKV infection, YFV infection or vaccination, combined DENV and ZIKV infection (DENV+ZIKV), ZIKV and YFV infection/vaccination (ZIKV+YFV), DENV and YFV infection/vaccination (YFV+DENV), or combined DENV, ZIKV, and YFV infection/vaccination (YFV+DENV+ZIKV), while an additional subset lacked detectable reactivity to any of the six orthoflaviviruses tested (Figs. S1A-D, G-H, Table S1). The specificity of viral protein identification was confirmed using monoclonal antibodies directed against E, prM, and NS1 proteins, which recognized the expected viral bands across orthoflavivirus lysates (Fig. S1E and S1F).

The study population ranged in age from 11 to 91 years (mean age, 42.1 years), with age-group distributions broadly reflecting the demographic composition of the Saúde population (Table S2). The male-to-female ratio was 1.24. Overall, serological evidence of prior orthoflavivirus exposure was highly prevalent within the cohort, with 213 participants exhibiting evidence of prior DENV infection, 61 prior ZIKV infection, and 175 prior YFV infection or vaccination, corresponding to seropositivity rates of 71.0%, 20.3%, and 58.3%, respectively (Fig. 1A and 1B). Fifty-nine individuals (19.7%) lacked detectable orthoflavivirus reactivity. Age-stratified analyses demonstrated relatively stable seroprevalence across age groups for DENV, ZIKV, and YFV exposures, without clear evidence of progressive accumulation with age (Fig. 1C). Consistent with previous epidemiological observations, DENV seropositivity was significantly higher among females than males (P = 0.02, Fisher’s exact test; Table S2).

**Figure 1.**
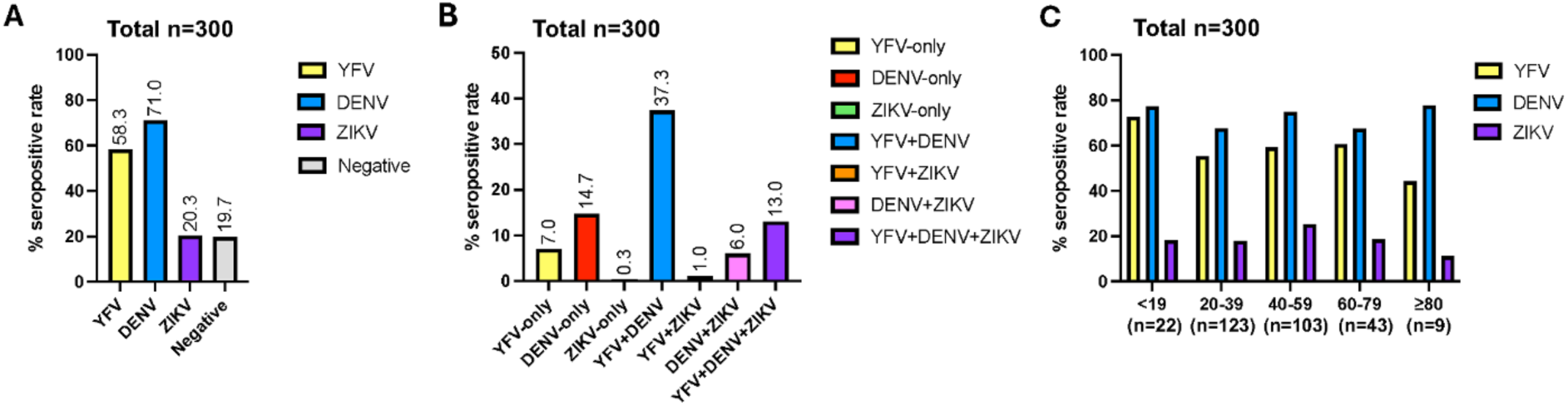
Orthoflavivirus seroprevalence in Saúde, Brazil determined by Western blot analysis. (A) Overall seropositive rates for yellow fever virus (YFV), dengue virus (DENV), and Zika virus (ZIKV), as well as seronegative individuals, among the study population (n=300). (B) Distribution of participants according to single or multiple orthoflavivirus-associated immune histories, including YFV-only, DENV-only, ZIKV-only, YFV+DENV, YFV+ZIKV, DENV+ZIKV, and YFV+DENV+ZIKV seropositivity. (C) Age-stratified seroprevalence of YFV, DENV, and ZIKV across study participants. Seropositivity was determined by Western blot detection of virus-specific anti-prM antibodies. Percentages above bars indicate the proportion of seropositive individuals within each category.

Notably, a substantial proportion of participants demonstrated serological evidence of multiple orthoflavivirus exposures, including DENV+ZIKV (6.0%), ZIKV+YFV (1.0%), YFV+DENV (37.3%), and YFV+DENV+ZIKV (13.0%) exposure profiles (Fig. 1A-B). The high frequency of individuals with overlapping orthoflavivirus immune histories provided a unique opportunity to investigate how multiple orthoflavivirus exposures shape antibody specificity, cross-reactivity, and functional activity in the context of pre-existing YFV immunity. To further interrogate these relationships, we selected representative groups for in-depth characterization, including YFV-only participants (n=9), whose immunity likely reflected either natural infection or vaccination, YFV+DENV participants (n=14), and YFV+DENV+ZIKV participants (n=12) from Saúde, together with plasma samples obtained from YFV-17D vaccinees in the United States (n=8).

Representative WB profiles from the distinct YFV-exposed groups are shown in Fig. 2. Among the YFV-only group, which included both YFV-17D vaccinees and individuals with presumed natural YFV exposure, anti-E antibodies exhibited broad cross-reactivity across all six orthoflavivirus lysates in most samples, whereas anti-prM responses remained restricted to YFV antigen (Fig. 2A-B; Fig. S1-2). In contrast, participants within the YFV+DENV group demonstrated extensive anti-E cross-reactivity across orthoflaviviruses, accompanied by broader anti-NS1 recognition patterns that frequently included DENV antigens and, in some cases, YFV or ZIKV antigens. Anti-prM antibodies in this group recognized DENV and YFV antigens, but not ZIKV or WNV antigens (Fig. 2C; Fig. S1-2). Participants with combined YFV+DENV+ZIKV exposure exhibited the broadest antibody recognition profiles, characterized by extensive cross-reactive anti-E and anti-NS1 responses across orthoflaviviruses and anti-prM recognition of DENV, YFV, and ZIKV antigens, while WNV reactivity was not found (Fig. 2D; Fig. S1-2). Collectively, these analyses revealed complex patterns of orthoflavivirus antibody cross-reactivity and antigen specificity across E, NS1, and prM proteins, reflecting the diverse immune histories present within this highly exposed population (Table S1).

**Figure 2.**
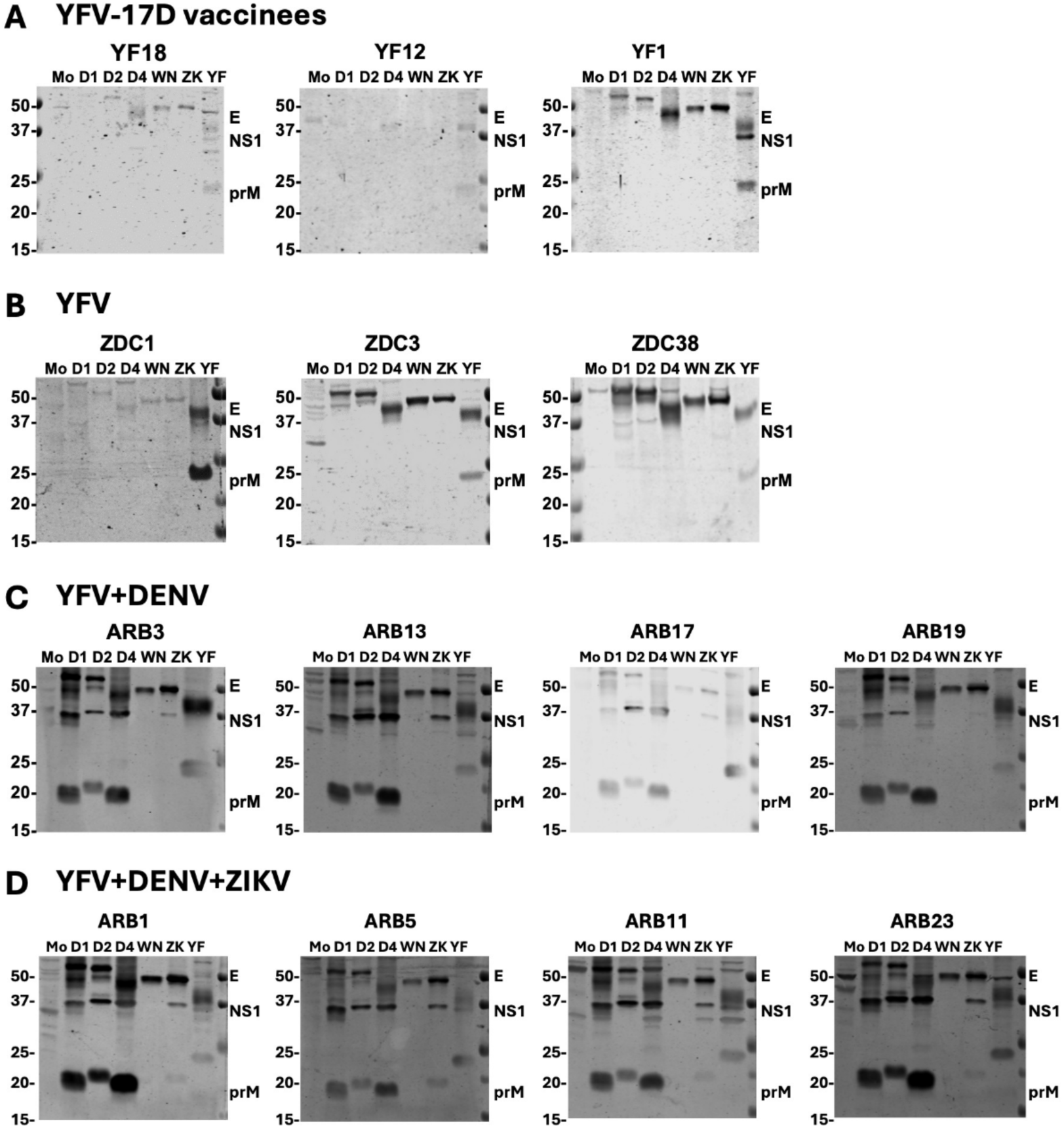
Antibody response to orthoflaviviruses in different YFV-exposed groups. Lysates derived from mock-, DENV1 (D1), DENV2 (D2), DENV4 (D4), WNV (WN), ZIKV (ZK), and YFV-17D (YF) infected Vero cells were subjected to SDS-12% polyacrylamide gel electrophoresis under non-reducing condition and Western blot analysis probed with plasma samples. Representative images of Western blot results from participants with previous (A) YFV-17D vaccinees (YF18, YF12, and YF1), (B) YFV infection or vaccination (ZDC1, ZDC3, and ZDC38), (C) YFV+DENV infections/vaccination (ARB3, ARB13, ARB17, and ARB19), and (D) YFV+DENV+ZIKV infections/vaccination (ARB1, ARB5, ARB11, and ARB23).

### Neutralizing antibody landscapes reflect cumulative orthoflavivirus exposure

Microneutralization assays were performed against YFV-17D, DENV1-4, ZIKV, and WNV to validate serological classifications and to define the breadth and specificity of neutralizing antibodies within each exposure group. Individuals in the YFV-only group exhibited strongly YFV-specific neutralization profiles. All YFV-17D vaccinees (8/8) displayed microneutralization 90 (NT_90_) titers ≥40 against YFV, confirming prior exposure history (Fig. 3A). However, only half (4/8) of YFV infection/vaccination group from Brazil had detectable NT_90_ titers against YFV, suggesting the possibility of infection by other genotypes such South America genotype I ^47^. In contrast, none of the samples in this group demonstrated detectable neutralization against DENV1-4, ZIKV, or WNV, despite considerable cross-reactive anti-E antibodies observed by Western blot. This dissociation between binding breadth and neutralization specificity is consistent with the notion that potent orthoflavivirus neutralization is largely mediated by type- or virus-specific E epitopes, whereas cross-reactive antibodies are often weakly neutralizing or non-neutralizing ^8^.

**Figure 3.**
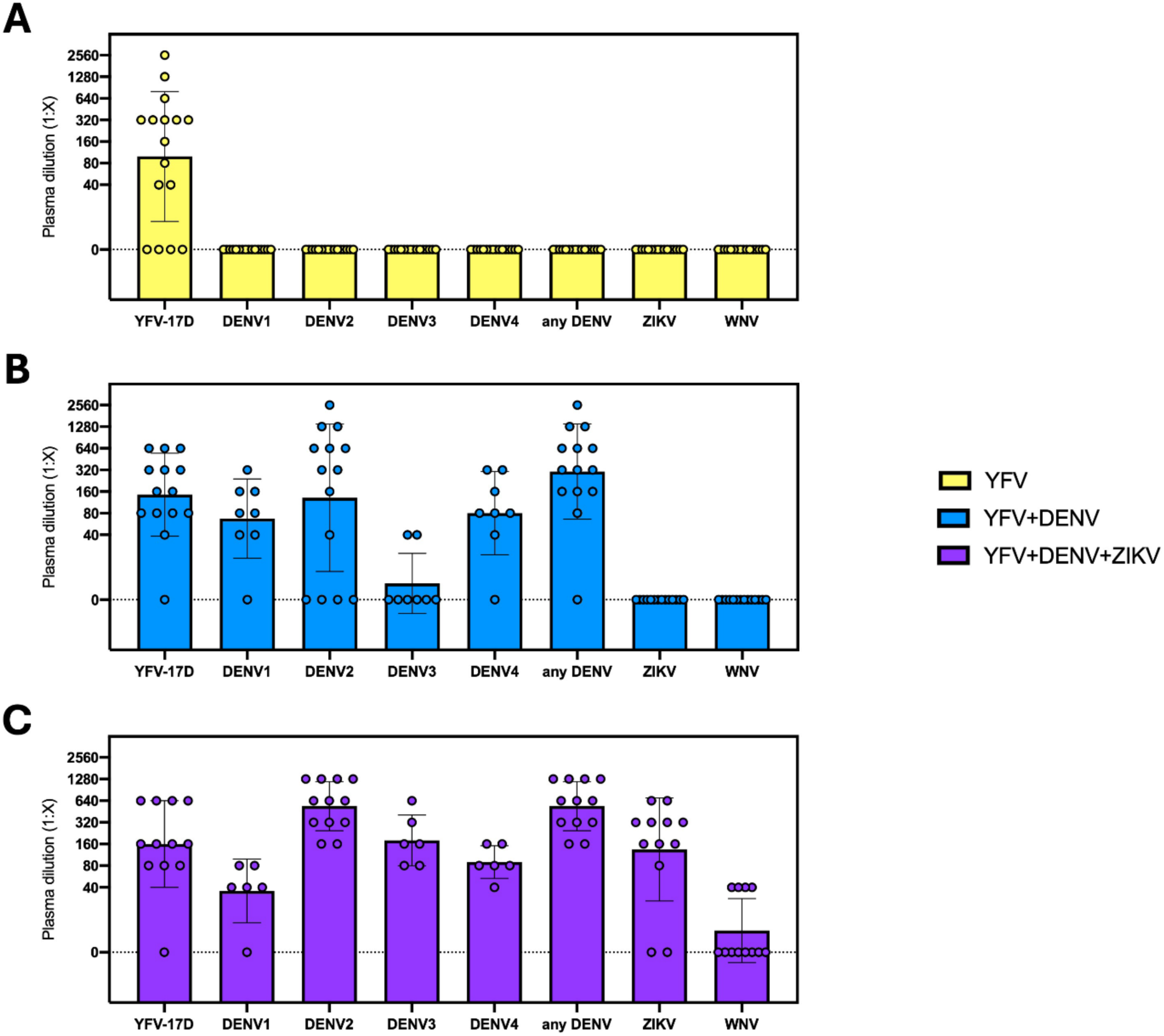
Neutralizing antibody profiles across orthoflavivirus exposure groups. Plasma from three groups, (A) YFV-only, (B) YFV+DENV, and (C) YFV+DENV+ZIKV, were evaluated for neutralizing antibody titers (NT_90_) against YFV-17D, dengue virus serotypes 1-4 (DENV1-DENV4), Zika virus (ZIKV), and West Nile virus (WNV). Plasma samples were serially diluted (starting from 1:40) and incubated with 100 PFU of virus prior to infection of Vero cells. The NT_90_ titer represents the reciprocal plasma dilution (1:X) at which 90% of Vero cells were protected from virus-induced cytopathic effects (CPE). Each symbol represents an individual plasma sample. NT_90_ titers <40 were designated as 0, which were shown along the dot lines.

Within the YFV+DENV group, neutralization breadth expanded substantially. A total of 92.9% (13/14) of samples neutralized YFV, confirming prior YFV exposure, while 71.4% (10/14) exhibited detectable neutralization against DENV2 (Fig. 3B). To further resolve serotype breadth, a subset of samples with available sera were tested across all DENV1-4. In this subset, neutralization was detected in 87.5% (7/8) of samples against DENV1, 71.4% (10/14) against DENV2, 25% (2/8) against DENV3, and 87.5% (7/8) against DENV4, resulting in detectable neutralization to any DENV serotype in 92.9% (13/14). Notably, 75% (6/8) of samples in this subset neutralized three or more DENV serotypes, suggesting either sequential heterotypic DENV infections or the accumulation of cross-neutralizing antibody responses typical of endemic transmission settings. Despite this expanded DENV neutralization breadth, no detectable neutralization was observed against ZIKV or WNV, supporting the serological classification based on anti-prM antibodies in this exposure group.

The YFV+DENV+ZIKV group exhibited the most complex and broadly reactive neutralization landscapes. In this group, 91.7% (11/12) of samples neutralized YFV-17D, 100% (12/12) neutralized any DENV, and 83.3% (10/12) neutralized ZIKV (Fig. 3C). In contrast to the YFV+DENV group, several samples, 33.3% (4/12) also demonstrated low level but measurable cross-neutralization against WNV despite no antibody reactivity to WNV by Western blot. These findings suggest the emergence of broadly cross-neutralizing antibody repertoires after multiple orthoflavivirus exposures spanning different serocomplexes. Collectively, these data indicate that increasing exposure complexity is associated with a progressive expansion of neutralization breadth, transitioning from virus-specific responses in the YFV-only group to multitypic DENV responses in the YFV+DENV group and ultimately to broadly cross-reactive neutralization profiles in the YFV+DENV+ZIKV group.

### Cumulative orthoflavivirus exposure increases in vitro enhancement potential

To determine whether the neutralization landscapes associated with each exposure profile translated into distinct functional enhancement phenotypes, we quantified in vitro ADE using FcγR-bearing U937 cells across YFV-17D, ZIKV, WNV, and DENV1-4. Serial plasma dilutions were tested to capture the canonical enhancement window that emerges under sub-neutralizing conditions.

Enhancement activity from the three groups were quantified as the area under the curve (AUC) against heterologous viruses YFV-17D, ZIKV, WNV and DENV1-4 (Fig. 4A). We additionally quantified the single highest peak fold enhancement (FE) to capture the the dilution at which the highest enhancement occurred (Fig. 4B). YFV-only plasma against YFV, ZIKV and WNV infections remained at or near baseline across the dilution series quantified as AUC or as the peak FE (Fig. 4A-B, Fig. S6A-C), with no consistent enhancement window observed. When extended to DENV1-4, YFV-only plasma again showed minimal enhancement for DENV1 and DENV2 (Fig. 4A-B, Fig. S6D-E), for both ADE parameters. Modest enhancement was detectable for DENV3, predominantly at higher dilutions (median 10000), and for DENV4 within a narrower intermediate dilution range (median 100) (Fig. S6F-G). Notably, DENV2 enhancement based on AUC or FE was significantly higher among those with anti-E antibodies cross-reactive to DENV2 than those without (Fig. S3A-B), underscoring the presence of cross-reactive binding antibodies contributed to ADE activity in the YFV-only group.

**Figure 4.**
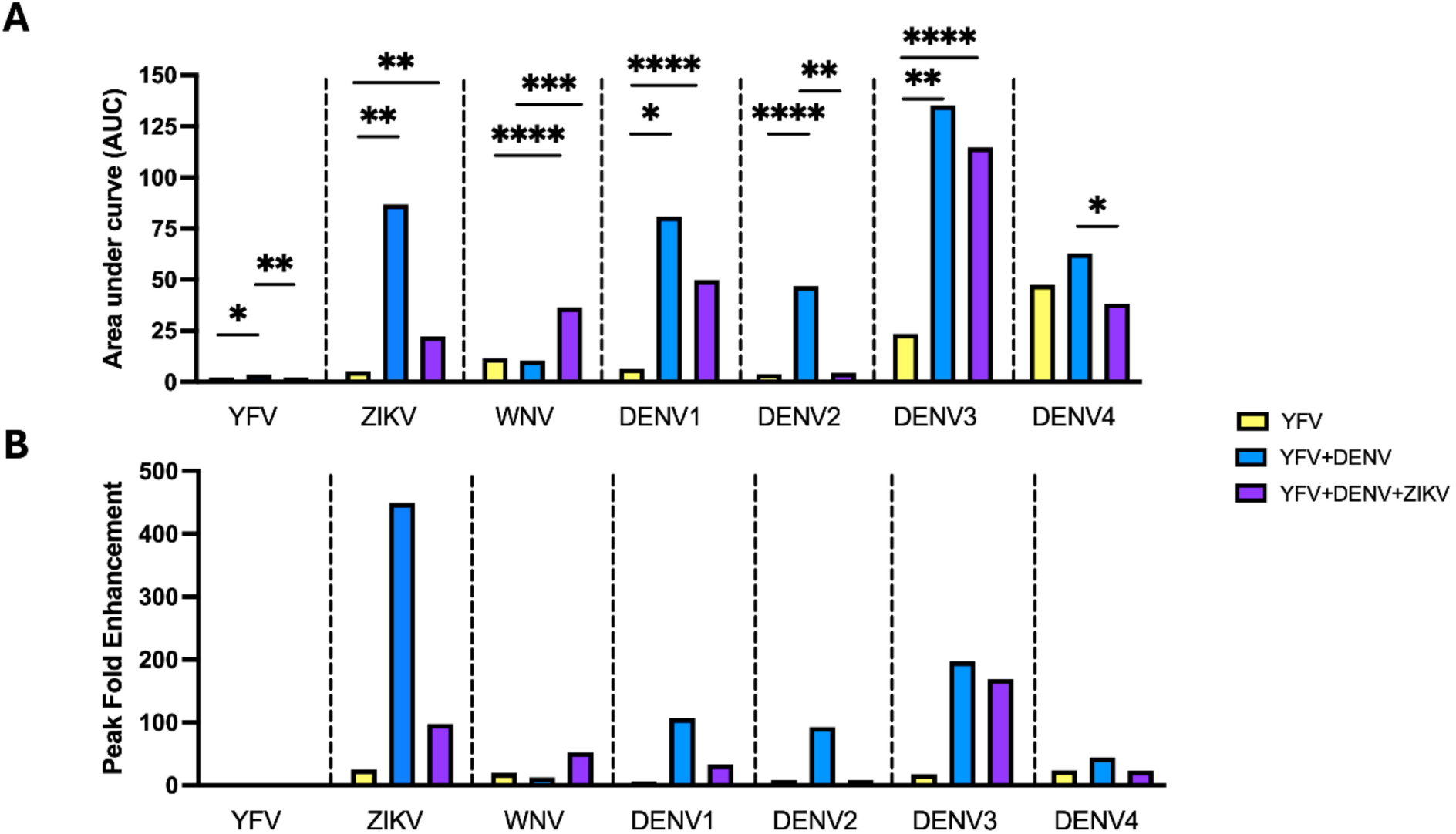
Peak fold enhancement and area under the curve values across orthoflavivirus exposure groups. Serially diluted plasma from three groups, YFV-only (yellow), YFV+DENV (blue), and YFV+DENV+ZIKV (purple), were evaluated for Fcγ receptor-mediated antibody-dependent enhancement (ADE) using U937 cells. Plasma samples were incubated with virus prior to infection of U937 cells, and infection was quantified by intracellular staining for orthoflavivirus envelope protein. (A) The area under the curve (AUC) across 5 dilutions (1:10-1:100,000) was quantified for all samples of each infection history and the mean was plotted against each virus. (B) The highest peak fold enhancement (FE) was plotted from each infection history against each virus. AUC comparisons between YFV vs YFV+DENV, YFV vs YFV+DENV+ZIKV and YFV+DENV vs YFV+DENV+ZIKV were calculated using two-tailed t-test. Significant *P* values are denoted with an asterisk (*), non-significant p values are not shown. Significant P values are as follow: *, P ≤ 0.05; **, P ≤0.01; ***, P ≤ 0.001, ****, P ≤ 0.0001.

In contrast, the YFV+DENV exposure group (Fig. 4A-B, S6H-N) exhibited clear and reproducible enhancement phenotypes across multiple heterologous viruses. ZIKV enhancement was most apparent within a discrete mid-to-high dilution window, with the group-average FE peaking around the 10^3^ reciprocal dilution range and reaching approximately ∼20-fold (Fig. S6I). YFV+DENV plasma demonstrated high AUC values across ZIKV and DENV1-4, with the highest AUC value against DENV3 (Fig. 4A).

Among the groups, YFV+DENV plasma displayed the highest peak FE to ZIKV across all samples and viruses tested against (Fig 4B). Among DENV1-4, enhancement was strongly serotype-dependent in both magnitude and dilution threshold. DENV3 demonstrated the highest peak FE, approaching ∼100-fold in the group average, with maximal activity occurring within an intermediate dilution range (Fig. S6M). DENV1, DENV2, and DENV4 exhibited intermediate enhancement amplitudes on the order of several tens-fold, with peaks occurring at similar or slightly shifted dilution windows (Fig. S6K, L, and N). These patterns align with the neutralization profiles in this cohort, in which DENV3 displayed comparatively lower neutralizing activity in a subset of samples, consistent with the established principle that ADE is most pronounced when antibody concentrations fall below the neutralization threshold and FcγR-mediated entry predominates.

The YFV+DENV+ZIKV group (Fig. 4A-B, S6O-U) displayed enhancement dynamics consistent with broader and higher-titer antibody repertoires in which the enhancement window shifts toward higher dilutions. AUC values in this group were significantly higher than the YFV-only group against ZIKV and WNV (Fig. 4A). Interestingly, AUC was high against DENV1, DENV3, and DENV4, but not DENV2 (Fig. 4A). Similar to the other exposure groups, YFV+DENV+ZIKV plasma resulted in no enhancement of YFV-17D (Fig. 4A-B, S6O), as well as minimal enhancement of DENV2 and DENV4 (Fig. 4A-B, S6S, U). However, modest enhancement was detectable for ZIKV (Fig. 4A-V, S6P), WNV (Fig. 4A-B, S6Q), and DENV1 (Fig. 4A-B, S6R), with substantial enahancement of DENV3 (Fig. 4A-B, S6T). Interestingly, for YFV-multiple exposed groups, the NT_90_ titers to DENV positively correlated with the dilutions at peak fold FE and inversely correlated with the peak FE or AUC considering all DENV serotypes together (Figs. S3C-E). Collectively, these in vitro data demonstrate that enhancement potential is strongly shaped by cumulative exposure history and varies by virus and serotype. Importantly, the dilution at which enhancement emerges shifts in accordance with neutralization breadth, supporting a model in which the net functional outcome reflects a dynamic balance between neutralizing and enhancing antibody fractions within polyclonal plasma.

### In vivo passive transfer demonstrates dose-dependent enhancement phenotypes to ZIKV and DENV2 challenges

To determine whether the in vitro enhancement phenotypes translated into physiologically relevant outcomes, plasma from each exposure group was passively transferred into *IFNAR1^-/-^* mice prior to challenge with either ZIKV or DENV2. This model enables direct evaluation of the functional consequences of human polyclonal antibody repertoires in vivo under conditions of impaired type I interferon signaling, which sensitizes mice to orthoflavivirus infection. To approximate sub-neutralizing versus neutralizing antibody conditions, two transfer volumes were used, including a low-dose transfer (2 µL) intended to model enhancing antibody concentrations and a high-dose transfer (100 µL) representing more protective, neutralizing conditions.

Across challenges, mice were monitored for viremia at early and later time points and for clinical outcomes including weight loss and survival.

Passive transfer of YFV-only plasma in the ZIKV lethal challenge model did not result in significant differences in ZIKV viremia at 3 dpi under either low-dose (Fig. 5A) or high-dose (Fig. 5B) conditions relative to naïve plasma controls. Viral loads declined similarly across groups, indicating that antibodies elicited by isolated YFV exposure do not measurably enhance early ZIKV replication in this model. Consistent with these virologic findings, survival kinetics were comparable between YFV-only plasma recipients and controls (Fig. 5C), and weight trajectories did not differ significantly between groups (Fig. 5D). Clinical scoring similarly demonstrated parallel disease progression across cohorts (Fig. S7B-C). Together, these data indicate that antibodies from individuals with isolated YFV exposure neither enhance ZIKV replication nor exacerbate disease severity under the conditions tested.

**Figure 5.**
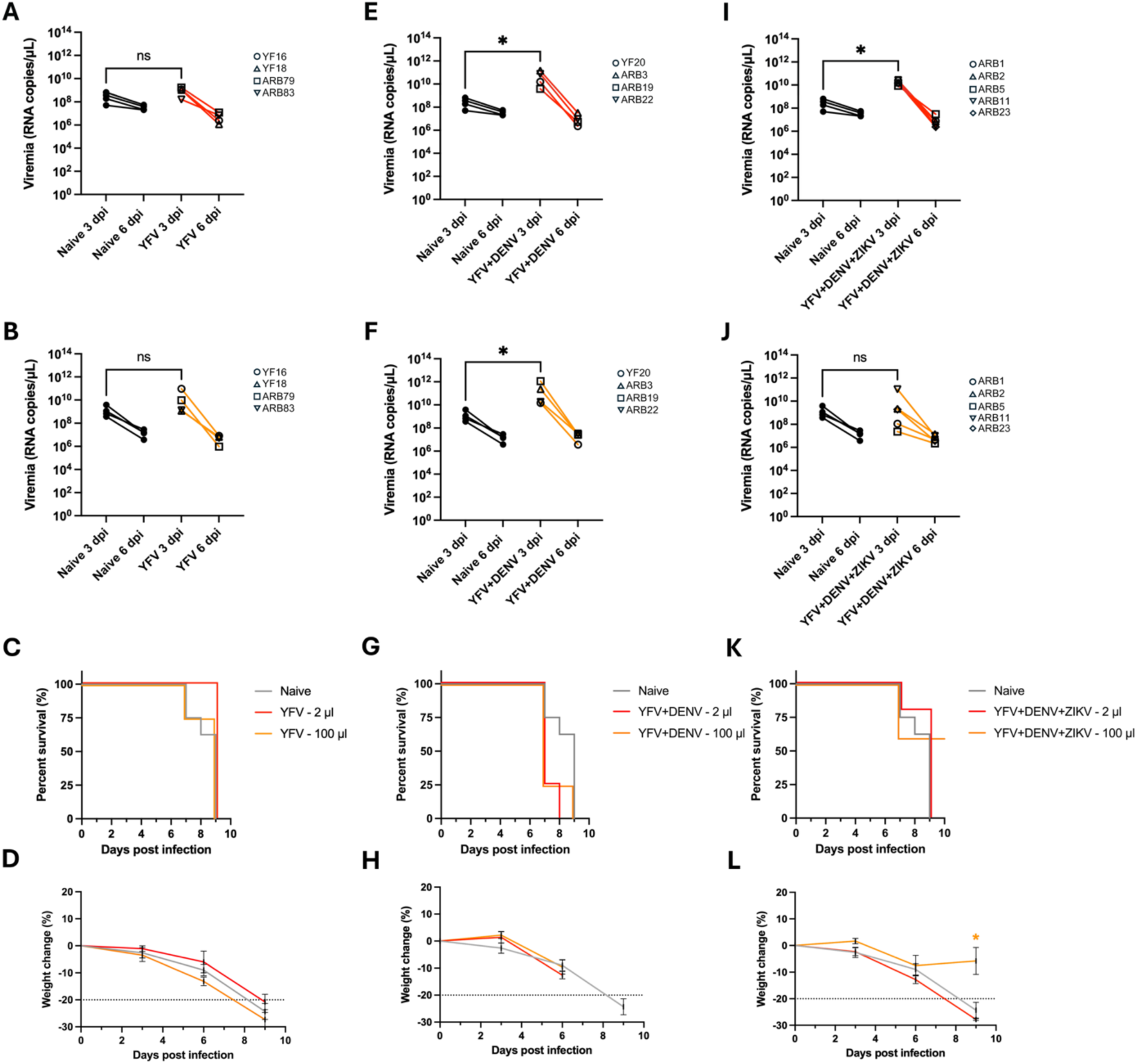
Viremia, survival, and weight change following passive transfer of plasma from distinct orthoflavivirus exposure groups in a lethal ZIKV challenge model. *IFNAR1^⁻/⁻^* mice received human plasma 24 hours prior to ZIKV challenge and were monitored for plasma viremia, survival, and weight loss. ZIKV viremia (RNA copies/µL) was quantified by RT-qPCR at 3 and 6 dpi. (A-B) Low-dose (2 µL) and high-dose (100 µL) transfer of YFV-only plasma. (C-D) Corresponding survival and weight change analyses. (E-F) Low-dose and high-dose transfer of YFV+DENV plasma. (G-H) Corresponding survival and weight change analyses. (I-J) Low-dose and high-dose transfer of YFV+DENV+ZIKV plasma. (K-L) Corresponding survival and weight change analyses. Individual symbols represent plasma from distinct human donors, with each point corresponding to an individual mouse. Gray lines indicate naïve plasma controls, whereas red and orange lines denote low-dose (2 µL) and high-dose (100 µL) plasma transfer, respectively. Viremia at 3 dpi was compared using Mann-Whitney U tests. Survival differences were analyzed by Kaplan-Meier analysis with Holm-Šídák multiple-comparisons testing. Weight change is shown as percent change from baseline body weight and was analyzed using two-tailed Student’s t-tests. *, P < 0.05; ns, not significant.

In contrast, plasma from YFV+DENV donors resulted in significantly elevated ZIKV viremia at 3 dpi under both low-dose (Fig. 5E) and high-dose (Fig. 5F) transfer conditions relative to naïve plasma controls. The increase in viral burden was observed across multiple donor samples and persisted despite the higher plasma transfer volume, indicating that antibody repertoires in this exposure group contain a substantial fraction capable of mediating in vivo enhancement. Concordant with increased early viral replication, survival kinetics were altered in mice receiving YFV+DENV plasma relative to naïve controls (Fig. 5G), with a trend in both the 2 and 100 µL groups of shorter time to mortaility, showing 3/4 (75%) of mice succumbing to infection by 7 dpi, compared to only 2/8 (25%) of naïve mice. Weight loss was more pronounced in plasma-transferred animals, in both the low-dose and high-dose conditions, with several mice reaching the predefined 20% weight-loss threshold by 7 dpi (Fig. 5H), a clinical end-point of the study. Clinical scoring demonstrated earlier onset of ruffled fur and hunched posture in YFV+DENV plasma recipients compared to controls (Fig. S7D-E). Collectively, these findings establish that sequential YFV+DENV exposure generates antibody repertoires capable of driving bona fide in vivo enhancement of ZIKV replication and disease.

Plasma from YFV+DENV+ZIKV donors demonstrated a concentration-dependent phenotype. Low-dose (2 µL) transfer resulted in significantly increased ZIKV viremia at 3 dpi relative to naïve plasma controls (Fig. 5I), consistent with infection occurring under sub-neutralizing conditions in which FcγR-mediated entry can predominate. In contrast, high-dose (100 µL) transfer did not produce a statistically significant increase in viremia at the group level (Fig. 5J), indicating that higher ZIKV antibody concentrations typically mitigate the enhancement phenotype observed at low dose. Survival outcomes were correspondingly improved relative to the low-dose condition, with an overall survival rate of 3/5 (60%), compared to the naïve plasma transfer group’s survival rate of 0/8 (0%). (Fig. 5K), and several animals demonstrated stabilization or recovery of weight during the observation period (Fig. 5L). Although inter-donor variability persisted, accelerated mortality was not observed at the group level under high-dose conditions compared to naïve controls.

We next examined in vivo enhancement in the DENV2 sublethal challenge model. In mice receiving plasma from individuals with YFV-only exposure, low-dose (2 µL) and high-dose (100 µL) passive transfers did not result in measurable enhancement of DENV2 replication. Across both experimental conditions (Fig. 6A-B), viremia levels at 2 dpi were comparable to naïve plasma controls, and viral loads declined similarly by 4 dpi. Although individual donor samples exhibited minor variability, no consistent upward shift in early viremia was observed at the group level, and comparisons with naïve controls were not significant. Consistent with the virologic data, survival kinetics were indistinguishable between YFV-only plasma recipients and naïve controls (Fig. 6C).

**Figure 6.**
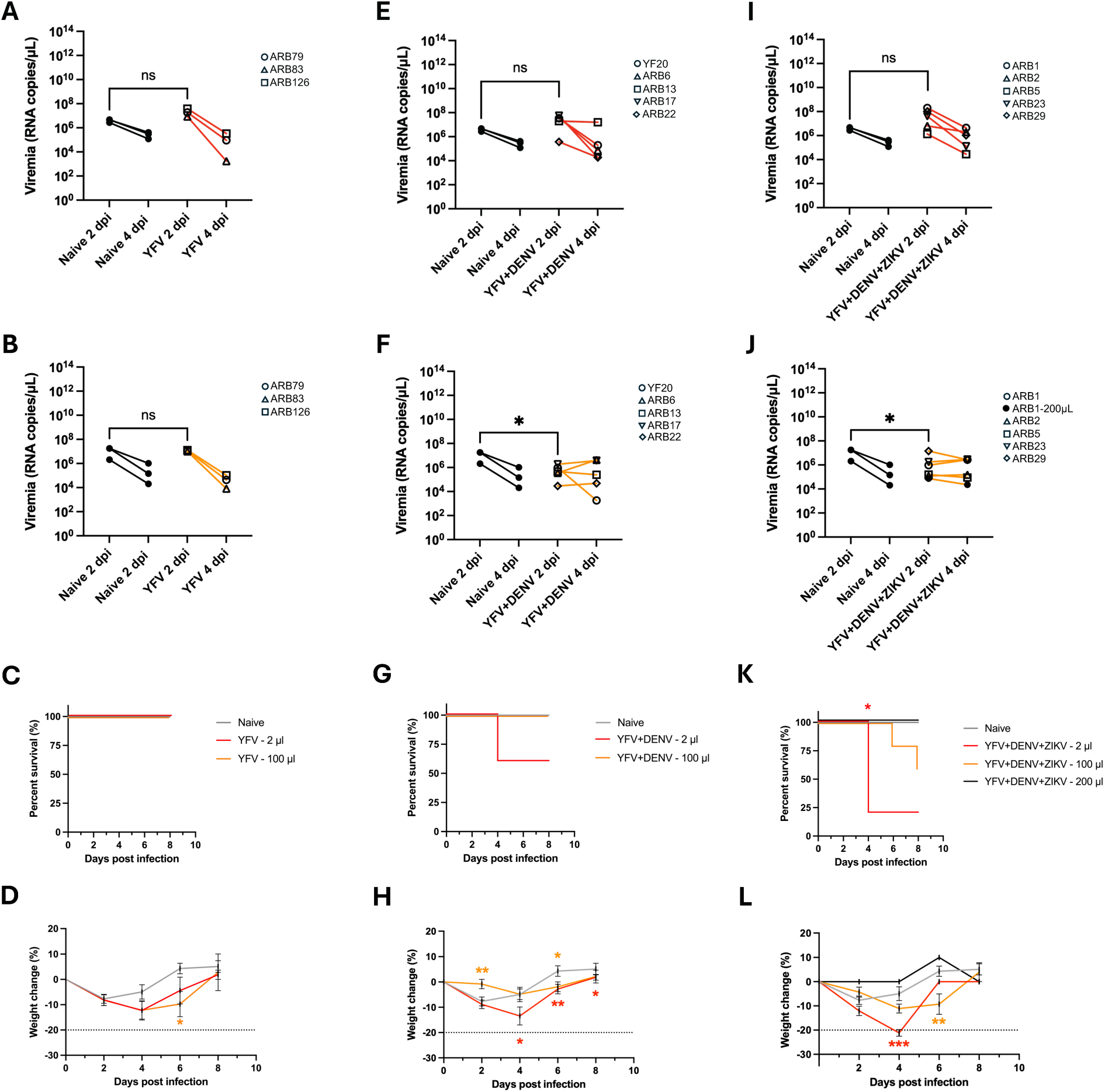
Viremia, survival, and weight change following passive transfer of plasma from distinct orthoflavivirus exposure groups in a DENV2 challenge model. *IFNAR1^⁻/⁻^* mice received human plasma 24 h prior to DENV2 challenge and were monitored for plasma viremia, survival, and weight loss. DENV2 viremia (RNA copies/µL) was quantified by RT-qPCR at 2 and 4 dpi. (A-B) Low-dose (2 µL) and high-dose (100 µL) transfer of YFV-only plasma. (C-D) Corresponding survival and weight change analyses. (E-F) Low-dose and high-dose transfer of YFV+DENV plasma. (G-H) Corresponding survival and weight change analyses. (I-J) Low-dose and high-dose transfer of YFV+DENV+ZIKV plasma. (K-L) Corresponding survival and weight change analyses, including 200 µL plasma transfer conditions where indicated. Individual symbols represent plasma from distinct human donors, with each point corresponding to an individual mouse. Gray lines indicate naïve plasma controls, whereas red and orange lines denote low-dose (2 µL) and high-dose (100 µL) plasma transfer, respectively; black lines indicate 200 µL plasma transfer. Viremia at 2 dpi was compared using Mann–Whitney U tests. Survival differences were analyzed by Kaplan–Meier analysis with Holm–Šídák multiple-comparisons testing. Weight change is shown as percent change from baseline body weight and was analyzed using two-tailed Student’s t-tests. *, P < 0.05; **, P < 0.01; ***, P < 0.001; ns, not significant.

Weight trajectories and clinical scores similarly demonstrated overlapping disease courses, with no evidence of accelerated symptom onset, increased weight loss, or earlier mortality in the 2 µL group, but the 100 µL group did have significantly increased weight loss at 6 dpi compared to naïve controls, but despite this change still recovered to pre-infection levels by 8 dpi (Fig. 6D, S8B-C). Collectively, these findings indicate that plasma from individuals with isolated YFV exposure does not measurably enhance DENV2 infection or disease severity in this model.

In contrast, plasma from YFV+DENV exposed individuals demonstrated a concentration-dependent shift in early viral kinetics following DENV2 challenge. At 2 dpi, low-dose (2 µL) transfer resulted in viremia levels that trended higher than naïve plasma controls (Fig. 6E), across 4/5 (80%) of samples, with little inter-donor variability. One sample (ARB22), displayed a substantial shift from the group average, starkly reducing viral load at 2 dpi compared to the naïve plasma transfer group. This pattern is consistent with infection occurring under sub-neutralizing antibody concentrations, in which FcγR-mediated viral entry can enhance early replication. However, when a higher plasma volume (100 µL) was transferred, the directionality of the effect reversed, with viremia at 2 dpi significantly lower relative to naïve controls (Fig. 6F), suggesting restoration of neutralization at increased antibody concentrations. By 4 dpi, viral loads declined across donor samples in both conditions, indicating that the early amplification observed under low-dose transfer was transient and did not produce sustained increases in viral burden. This early kinetic shift was reflected in survival outcomes, with survival curves showing 40% (2/5) succumbing to infection by 8 dpi, compared to 100% survival in the naïve plasma transfer group (Fig. 6G). Within this group, the mouse receiving plasma from YF20 died at 4 dpi. The mice receiving plasma from ARB13, which showed persistently high levels of viremia (Fig. 6E), also succumbed to infection by 8 dpi. Similarly, weight change trajectories between the low-dose transfer group were significantly lower at 4, 6 and 8 dpi (Fig. 6H). Inversely, the high-dose transfer group had significantly increased weight gain at 2, yet transient weight loss at 6 dpi, compared to naïve controls (Fig. 6H). Clinical scoring likewise showed earlier onset of ruffled fur and hunched posture in a subset of animals receiving YFV+DENV plasma (Fig. S8D-E). Together, these data support a concentration-dependent enhancement phenotype in plasma from sequential YFV+DENV exposure histories, characterized by increased early DENV2 replication under sub-neutralizing conditions and attenuation of viremia at higher antibody concentrations. The transient nature of the effect suggests that antibody-mediated modulation is most pronounced during the initial phase of viral expansion.

In the YFV+DENV+ZIKV exposure group, passive transfer revealed more heterogeneous outcomes across donor samples, consistent with the broader and more complex neutralizing antibody landscapes observed in this group. Low dose transfer conditions trended towards increased DENV2 viremia relative to naïve plasma controls, consistent with enhancement occurring with transfer of ZIKV cross-reactive antibodies to DENV2 (Fig. 6I). In contrast, the high dose transfer conditions significantly decreased DENV2 viremia (Fig. 6J). These enhancement profiles were reflected in survival curves, with the low-dose transfer group experiencing significantly earlier mortality at 4 dpi (Fig. 6K). The 100 μL high-dose transfers demonstrated non-signficant increased mortaility compared with naïve group, with 33% (2/6) of mice succumbing to infection by 8 dpi (Fig. 6K). However, a 200 μL high-dose transfer provided complete protection against earlier time to mortality (Fig. 6K). Weight trajectories and clinical scoring similarly demonstrated dose-dependent variability, with the low-dose transfer group animals exhibiting significantly more pronounced weight loss and symptom progression at 4 dpi (Fig. 6L, Fig. S8G). The high-dose groups showed significantly decreased weight loss at 6 dpi, but the surviving 66% (4/6) of animals were able to recover to pre-infection weights (Fig. 6L). This dose-dependent variability is consistent with differences in neutralizing antibody titers and functional antibody composition across individuals with complex exposure histories. In this group, the polyclonal plasma likely contains both strongly neutralizing and enhancing antibody fractions, such that the net in vivo outcome depends on the relative abundance of these fractions at a given antibody concentration.

## Discussion

ADE is a central immunopathogenic phenomenon in dengue and a persistent concern across orthoflaviviruses, yet the extent to which ADE manifests depends on the composition and concentration of antibody repertoires sculpted by multiple exposures ^48,49^. This question is particularly relevant in regions where YFV infection or YFV-17D vaccination overlaps with endemic ZIKV and DENV circulation, and where extensive serologic cross-reactivity obscures prior exposure histories. By combining serocomplex-specific anti-prM serology with microneutralization, FcγR-dependent in vitro enhancement assays, and in vivo passive-transfer challenge, this study defines how YFV-associated immune histories shape the balance between neutralizing and enhancing antibody functions. The data support three overarching conclusions. First, isolated YFV exposure generates robust YFV-neutralizing activity but does not yield a dominant antibody fraction that measurably enhances heterologous orthoflavivirus infection. Second, sequential exposure incorporating ZIKV and/or DENV broadens neutralization but also increases FcγR-dependent enhancement potential in vitro. Third, in vivo phenotypes are strongly concentration-dependent, consistent with canonical ADE kinetics in which enhancement emerges under low antibody levels or sub-neutralizing conditions and is typically mitigated as antibody levels increase ^15,50,51^.

A defining observation is the dissociation between broad cross-reactivity and functional neutralization breadth. Across exposure groups, WB reactivity to E was widely cross-reactive, whereas neutralization in the YFV-only group remained virus-specific, with no detectable neutralization of DENV1-4, ZIKV, or WNV. This pattern is consistent with several previous studies demonstrating that the most potent orthoflavivirus neutralizing antibodies primarily recognize serotype- or virus-specific epitopes, often quaternary epitopes on E, whereas cross-reactive antibodies commonly target conserved determinants that bind efficiently but neutralize weakly or not at all ^8,52^. Thus, binding breadth should not be interpreted as functional breadth, particularly in populations with layered flavivirus immunity.

A major barrier to studying multiple orthoflavivirus immunity is reconstructing exposure history in endemic settings. Anti-prM antibodies provided greater serocomplex resolution than E or NS1 binding and enabled stratification into YFV-only, YFV+DENV, and YFV+DENV+ZIKV profiles that were independently verified by microneutralization test. The utility of this approach is conceptual as well as practical, as ADE is a conditional phenotype that cannot be meaningfully interpreted without knowing whether antibody repertoires reflect single exposures or repeated immunologic recall and reshaping. Serocomplex-resolved strategies therefore provide a tractable framework for linking immune history to functional outcomes in settings where conventional serology is confounded by cross-reactivity ^46,53^.

In vitro ADE assays using FcγR-bearing U937 cells indicated that cumulative orthoflavivirus exposure increases enhancement potential, but in a virus- and serotype-dependent manner that aligns with the interplay between neutralization potency and cross-reactive binding. Plasma from YFV-only individuals showed minimal enhancement across heterologous viruses, whereas multiple exposure groups exhibited robust enhancement, including pronounced activity against ZIKV and multiple DENV serotypes. These findings are consistent with the established principle that enhancement peaks within a discrete enhancement window at antibody concentrations below the neutralization threshold but sufficient to opsonize virions for FcγR-mediated entry ^15^. Reviews of dengue ADE also emphasize that enhancement includes both “extrinsic” effects (increased entry into FcγR-expressing cells) and “intrinsic” effects (FcγR signaling that can modulate antiviral responses), reinforcing that both entry and downstream pathways can contribute to increased effective replication under sub-neutralizing conditions ^54^.

Sequential exposure likely increases enhancement potential by expanding cross-reactive antibody lineages targeting conserved epitopes, including epitopes in E domain II such as the fusion loop, which is strongly associated with broad binding and variable neutralization ^8,24,52,55,56^. A relevant precedent is the demonstration that pre-existing tick-borne encephalitis immunity can skew responses to YFV-17D toward broadly cross-reactive antibodies with increased in vitro ADE potential against ZIKV and DENV ^45^, supporting the notion that immune history can bias the repertoire toward conserved, potentially enhancing specificities. The present data extend this concept from sequential vaccination paradigms to endemic, multi-exposure settings, where infections and vaccination together shape antibody function.

In vivo passive-transfer experiments sharpen these inferences by directly testing physiological consequences and antibody concentration effects. Following ZIKV challenge, YFV-only plasma did not significantly increase viremia at either transfer dose and did not alter survival, weight loss, or clinical scores relative to controls, indicating that isolated YFV exposure does not readily generate an enhancing antibody fraction detectable in this model. In contrast, YFV+DENV plasma significantly increased ZIKV viremia at 3 dpi following both low- and high-dose transfer, and was accompanied by faster time to mortality. This phenotype suggests that early virologic amplification can increase mortality without necessarily shifting kinetics in weight change and clinical scores. Importantly, the YFV+DENV and YFV+DENV+ZIKV groups exhibited a classic titer-dependent pattern, especially with the low-dose transfers increasing ZIKV viremia, and with the most severe clinical course linked to the plasma sample associated with the highest viremia. The YFV+DENV+ZIKV high-dose transfers abrogated the viremia signal and was associated with preserved survival, though not achieving complete protection, probably due the neutralizing antibodies against ZIKV were below the threshold neutralization titers of protection, which was estimated to be 1:25 at 2 hours before challenge based on a recent study with a similar model ^29^; our estimated NT titers were 1:3 to 1:6 at 24 hours before challenge. This dose dependence is a hallmark of ADE and supports a model in which sequential exposure produces repertoires containing both enhancing and neutralizing fractions whose net effect is determined by antibody concentration ^15,57^. Interestingly, a significant correlation was found between relative ZIKV viremia in vivo and the FE of ZIKV in vitro for the low dose transfer or both low and high dose transfers together (Spearman correlation coefficient r=0.5750, *P*=0.04 or r=0.4648, *P*=0.02, respectively), but not the AUC or peak FE, suggesting the predictive value of in vitro ADE at specific concentrations (relevant to in vivo concentrations) rather than overall ADE activities (Fig. S4).

The DENV2 passive-transfer results further support the concentration-dependent model observed with ZIKV challenge. DENV2 is frequently used in murine enhancement studies because reproducible infection kinetics and quantifiable viremia enable detection of both enhancing and protective effects across antibody doses ^24,28,58^. In multiple exposure groups, modulation of viral kinetics was most apparent at the early 2 dpi timepoint, where low-dose plasma transfer trended toward increased viremia (YFV+DENV and YFV+DENV+ZIKV), while high-dose transfer shifted directionally toward reduced viral burden (YFV+DENV) though not to 100% survival, probably due to the neutralizing antibodies against DENV2 that did not reach the threshold neutralization titers of protection, which was estimated to be 1:400 before challenge based on a DENV2 in AG129 model ^24^. Although these comparisons approached but did not uniformly achieve significance prebably related to small sample size, the consistent bidirectional pattern across antibody concentrations is mechanistically consistent with classical ADE kinetics. Under sub-neutralizing conditions, cross-reactive antibodies can amplify early viral expansion through FcγR-mediated entry, whereas higher antibody concentrations restore neutralization and restrict viral replication. The fact that these effects were largely confined to early time points suggests that antibody-mediated modulation primarily influences the initial phase of systemic viral dissemination. Prior studies demonstrating heterologous orthoflavivirus antibody-mediated enhancement of DENV infection provide a conceptual framework for these findings and reinforce the biological plausibility of concentration-dependent ADE ^14,24,30,50^. Notably, the magnitude of the in vivo phenotype differed between ZIKV and DENV2 challenge, with more robust enhancement observed following ZIKV infection, which correlated significantly with an ADE parameter in vitro (Fig. S4), whereas no correlation between DENV2 phenotypes in vivo and ADE activities in vitro (Fig. S5). This likely reflects differences in baseline neutralization titers within each exposure group, serocomplex relatedness between challenge virus and priming exposures, and virus-specific replication kinetics in the *IFNAR1^⁻/⁻^* model.

Several considerations should be incorporated to contextualize interpretation. First, multitypic neutralization patterns cannot fully resolve infection order; thus, the exposure groups represent cumulative complexity rather than precise temporal sequences. Second, *IFNAR1^-/-^* mice provide a sensitized system for detecting antibody-mediated effects in particular ADE, but type I interferon impairment does not recapitulate intact human innate control; the model is therefore best positioned for mechanistic studies rather than direct quantitative inference to human severity. Third, FcγR repertoires differ between mice and humans ^59^, and human IgG subclass distributions and Fc glycosylation can further modulate FcγR engagement ^60^, contributing to the known heterogeneity of ADE across individuals and experimental systems. Finally, serotype- and strain-specific features can influence enhancement windows ; accordingly, conclusions are most appropriately framed around immune history and concentration dependence rather than asserting universal serotype equivalence.

Overall, these findings support a framework for YFV-associated immune histories in endemic settings. Isolated YFV exposure produces strongly YFV-focused neutralization and does not, on its own, establish a dominant enhancing antibody fraction against ZIKV or DENV2 in vivo. However, as sequential exposures accrue, particularly incorporating DENV and ZIKV, neutralization breadth expands alongside an increased propensity for FcγR-dependent enhancement that becomes apparent under sub-neutralizing conditions. These results reconcile why YFV immunity is often epidemiologically neutral with respect to heterologous severity while still participating in enhancement phenotypes after additional orthoflavivirus exposures reshape the repertoire ^36,61^. From a public health and vaccinology perspective, the data argue against presuming that YFV immunity alone elevates ADE risk, but emphasize that layered orthoflavivirus immunity can generate concentration-dependent risk windows that may become clinically relevant as antibody titers wane or as individuals encounter heterologous viruses at specific points along their immune trajectories. Future studies should focus on deconvulating the direct immunological impact of recalling pre-existing YFV immunity to subsequent heterologous orthoflavivirus infections.

## Materials and Methods

### Human samples

This study was approved by the Institutional Review Board (IRB) of the University of Hawaii (2019-00910 and 2021-00947). A total of 300 participants from Saúde, a town with a population of ∼11,000 in Bahia, a Northeastern state in Brazil, were enrolled with informed consents between June 2021 and February 2022. All residents were invited by the local government to participate in an arbovirus serological study at local health centers or schools. To ensure all 6 areas in Saúde were covered, house visit was conducted to recruit additional participants in areas where less than 50 participants were initially enrolled. Questionnaires including basic demographic information and blood sampling were obtained. The study was approved by the Comitê de Ética em Pesquisa da Maternidade Climério de Oliveira/UFBA, Brazil (CAAE: 25336819.3.

0000.5543/4.691.233, 2019). Samples of YFV-17D vaccinees were from the US (n=10) based on history of YFV-17D vaccination; two had previous DENV infection as verified by WB or mocroneutralization assay (7.59). This study was determined to be exempt by the University of Hawaii and Rutgers Robert Wood Johnson Medical School IRBs.

### Vero cell lysate and WB analysis

Vero cells and Vero cells infected with DENV1 (Hawaii strain), DENV2 (NGC strain), DENV4 (H241 strain), ZIKV (PRVABC59 strain), WNV (NY99 strain), YFV-17D (vaccine strain) or uninfected (mock) were lysed with NP-40 lysis buffer (1% NP-40, 50 mM Tris pH 8.0, 150 mM NaCl, 2 mM EDTA, and 1 mM Na_3_VO_4_) when 50% of cells were found to have cytopathic effects. The cell lysates were subjected to SDS-12% polyacrylamide gel electrophoresis under non-reducing condition (2% SDS, 0.5 M Tris pH 6.8, 20 % glycerol, 0.001 % bromophenol blue, final) (59), followed by transfer to nitrocellulose membrane (Trans-Blot Turbo RTA Midi Transfer Kit, BioRad), hybridization with human serum/plasma samples (1:200 dilution) or mouse monoclonal antibody and secondary antibody (IRDye® 800CW-conjugated goat anti-human or anti-mouse IgG at 1:10000). The signals were detected by Li Cor Odyssey classic (LiCor Biosciences) and analyzed by Image Studio software (59,60).

### Cells

Vero cells (kidney epithelial cells derived from a normal, adult African green monkey, *ATCC, Cat#: CCL-81.4*) were grown in EMEM media containing Earle’s Balanced Salt Solution, non-essential amino acids (NEAA), 2mM L-glutamine, 1 mM sodium pyruvate, 1500mg/L sodium bicarbonate and heat-inactivated 10% fetal bovine serum (FBS) and 1% penicillin-streptomycin solution. U937 cells (pro-monocytic human cell line isolated from a patient with histiocytic lymphoma; *ATCC, Cat#: CRL-1593.2*) were grown in RPMI-1640 media with L-glutamine, 10% FBS and 1% 1% penicillin-streptomycin solution. All cell lines with grown in an incubator at 37°C with 5% CO_2_.

### Virus production and tittering for in vitro and in vivo assays

YFV-17D, ZIKV (PRVABC59), and WNV (Bird 114) were all obtained from BEI resources and subsequently propagated. DENV1 (TH-Sman), DENV3 (SL 5-29-04) and DENV-4 (UIS 497) were all obtained from BEI Resources. DENV2 (S221), was kindly provided by Dr. Sujan Shresta from La Jolla Institute for Immunology. In brief, all viruses were propagated in Vero cells, and the viral titers were determined utilizing the TCID_50_ assay as previously described ^62^. The TCID_50_/mL value was used to calculate the plaque forming units (PFU) by using a conversion factor of 0.7 as previously described^58^.

### Microneutralization assay

We used previously described methods for the 96 well plate microneutralization assay ^37^. Vero cells were seeded at a density of 1 x 10^4^ one day prior to performing the assay. Cells were checked under the microscope 24 hours post attachment for 80% confluency. Plasma samples were diluted two-fold from 1:40 to 1:2560 in EMEM media supplemented with 2% FBS and L-Glutamine. In a 96 well plate, 100 PFU of YFV-17D, DENV1-4, ZIKV and WNV was added to diluted plasma samples, and the covered plate was incubated for 1 hour at 37°C in a 5% CO_2_ incubator. After incubation, plasma-virus complexes were added to adherent Vero cells and left to incubate for 5 days for YFV-17D, ZIKV and WNV and 7-10 days for DENV1-4. Cytopathic effects (CPE) were monitored via positive controls (orthoflavivirus, cells and media) until 100% cell CPE was achieved. In addition, 2 negative controls per plate (only cells and media) were added to ensure cell viability in the absence of orthoflavivirus.

### In vitro antibody-dependent enhancement (ADE) assay

We used previously described methods for the ADE assay. Briefly, in RPMI-1640 media with 2% FBS, plasma samples were serially diluted 10-fold from 1:10-1:100,000. Plasma samples were incubated for one hour with a multiplicity of infection of 1, 5, 2, and 0.5 MOI for YFV-17D, DENV1-4, ZIKV and WNV, respectively for one hour at 37°C in a 5% CO2 incubator. After incubation, U937 cells were resuspended for 2 hours with the plasma-virus mixture at 37°C in a 5% CO2 incubator. Cells were left to incubate for 2 days for YFV-17D, 4 days for DENV1-4 and ZIKV, and 1 day for WNV. Plasma-virus mixture supernatant was removed, and cells were stained for viability staining (live-dead red), fixed in paraformaldehyde and stained intracellularly with 4G2-Alexa Flour 532 Novus Biologicals, Cat# NBP2-52666AF532); a pan-orthoflavivirus envelope (E) targeting antibody. 2 x 10^4^ cells were acquired by the Cytek Aurora instrument to characterize a population of cells that stained positive for 4G2-Alexa Fluor 532, representing cells infected via Fc*γ*R-mediated antibody-dependent enhancement. Furthermore, flow cytometry analysis was performed using FlowJo software version 10.10.0.

### In vivo antibody-dependent enhancement

For characterization into the potential ADE effects of plasma *in vivo*, plasma samples with high FE *in vitro* were selected for further testing. We transferred 2 µL and 100 µL volumes of plasma intraperitoneally into five-week-old IFNAR1^-/-^ mice. Twenty-four hours post plasma transfer, mice were challenged retro-orbitally with 1 x 10^3^ PFU of ZIKV (PRVABC59) and 2 x 10^7^ PFU of DENV2 (S221). Submandibular blood samples were collected 3 and 6 days-post infection for ZIKV and 2 and 4 days-post infection (dpi) for DENV-2 to assess viremia after passive immunization. Additionally, mice were monitored daily for weight loss and clinical symptoms of ZIKV or DENV2 disease progression. Survival end points were recorded until 9 dpi for ZIKV with YFV and YFV+DENV plasma transfers. Survival was recorded until 13 dpi for ZIKV with YFV+DENV+ZIKV transfers. All survival endpoints were recorded until 8 dpi for DENV2 across plasma transfers groups.

### Clinical scoring of ZIKV and DENV2 disease

Assessment of both ZIKV and DENV2 pathogenesis in the IFNAR1^-/-^ mice were scored on a 0-5-point scale. Clinical symptoms include: 0, healthy, no symptoms; 1, ruffled fur; 2, ruffled fur and hunched posture; 3, lethargy or paralysis; 4, death.

### RNA extraction and one-step Taq man reverse transcription and real time polymerase chain reaction

Viral RNA was extracted from mouse plasma samples after ZIKV and DENV2 infection were done following the instructions from the QIAamp Viral RNA Mini Kit (QIAGEN). In brief, 10 µL of plasma was diluted in 130 µL of PBS. Samples were eluted in 60 µL of RNase free water. Extracted RNA was subject to one-step reverse transcription and amplified in via PCR with a master mix and primer-probe mixture from Integrated DNA Technology (IDT). The QuantStudio 3 was used to thermal cycle at the following conditions: reverse transcription, 50°C for 15 minutes; polymerase activation, 95°C for 3 minutes; denaturation, 95°C for 15 seconds; annealing and extension, 60°C for 1 minute, for 40 amplification cycles. The primers for ZIKV were used from a previous study ^63^: Forward primer, 5′ (CCGCTGCCCAACACAAG) 3’; Reverse primer, 3’ (TACAGACGTTTTCTTGCAATCACC) 5’ and probe, (5’-FAM-AGCCTACCTTGACAAGCAGTCAGACACTCAA-3’).The primers used for DENV-2 amplification were used from a previous publication ^64^; Forward primer, 5′ (GCAGAAACACAACATGGAACRATAGT) 3′; Reverse primer, 3′ (TGATGTAGCTGTCTCCRAATGG 5′) and probe (5′-FAM-

TCAACATAGAAGCAGAACC-TAMRA-3′). ZIKV and DENV-2 standard curves were generated utilizing genomic RNA from BEI resources. The number of ZIKV and DENV2 RNA copies was calculated using the genome equivalents formula: [ssRNA concentration (g/µL) * 6.022 × 10^23^(copy/mol)]/ [ssRNA length * 340 (g/mol)]. A stock solution of 10^8^ RNA copies/µL was prepared and serially diluted 10-fold to generate working solutions ranging from 10^0^ to 10^7^ copies/µL.

### Statistics

Neutralization titers were analyzed on log-transformed data where appropriate, and geometric mean titers were calculated for group-level comparisons. For in vitro ADE assays, fold-enhancement values were calculated relative to virus-only controls at each dilution, as previously described ^58^. For in vivo experiments, viremia levels at the 2 or 3 dpi time point were compared between the two independent groups (YFV-only versus YFV+DENV or YFV+DENV+ZIKV) using Mann-Whitney U tests (for nonparametric data). Survial differences between groups were assessed by the Kaplan-Meier survival analysis followed by followed by Holm-Šídák’s multiple comparisons test. *P* values less than 0.05 were considered statistically significant. Non-significant differences were not indicated. Weight loss trajectories between two or more groups over time were analyzed using two-sided student’s t-test. For all tests p values <0.05 were considered statistically significant. Any non-signifcant *P* value was not shown on the graph. All statistical analyses were performed using GraphPad Prism version 10.

## Data Availability

All data supporting the findings of this study are available within the article and its supplemental information.

## Acknowledgements

We would like to thank Rutgers Global Health Institute, Rutgers Robert Wood Johnson Medical School, and The Child Health Institute of New Jersey for their continued support. This study was funded in part by R01AI86073 to WKW and BBH.

## Supplementary Tables

**Supplementary Table S1.**
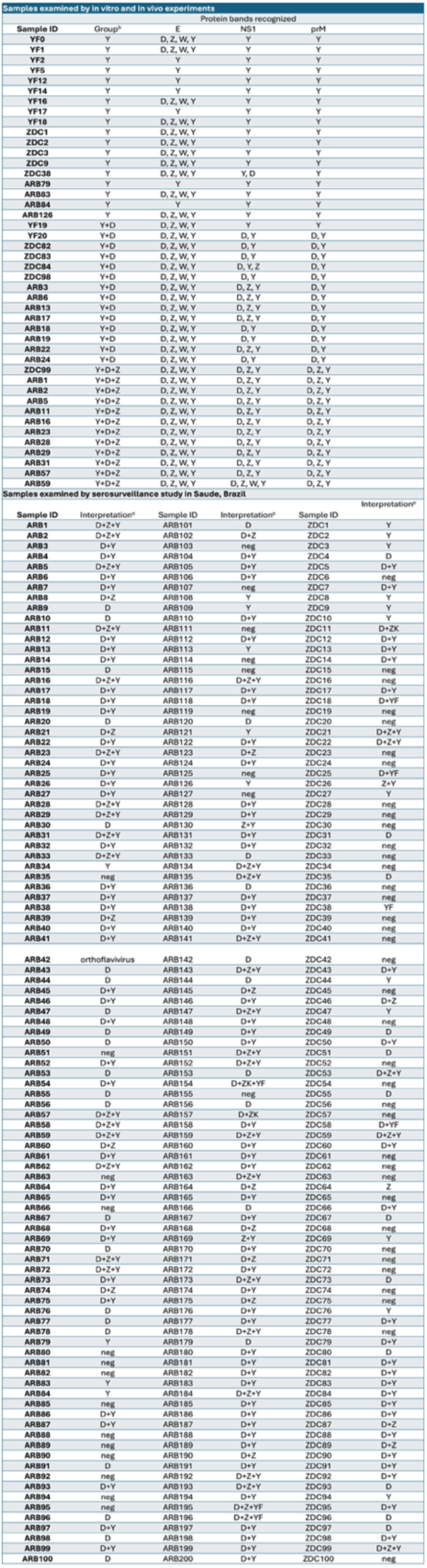
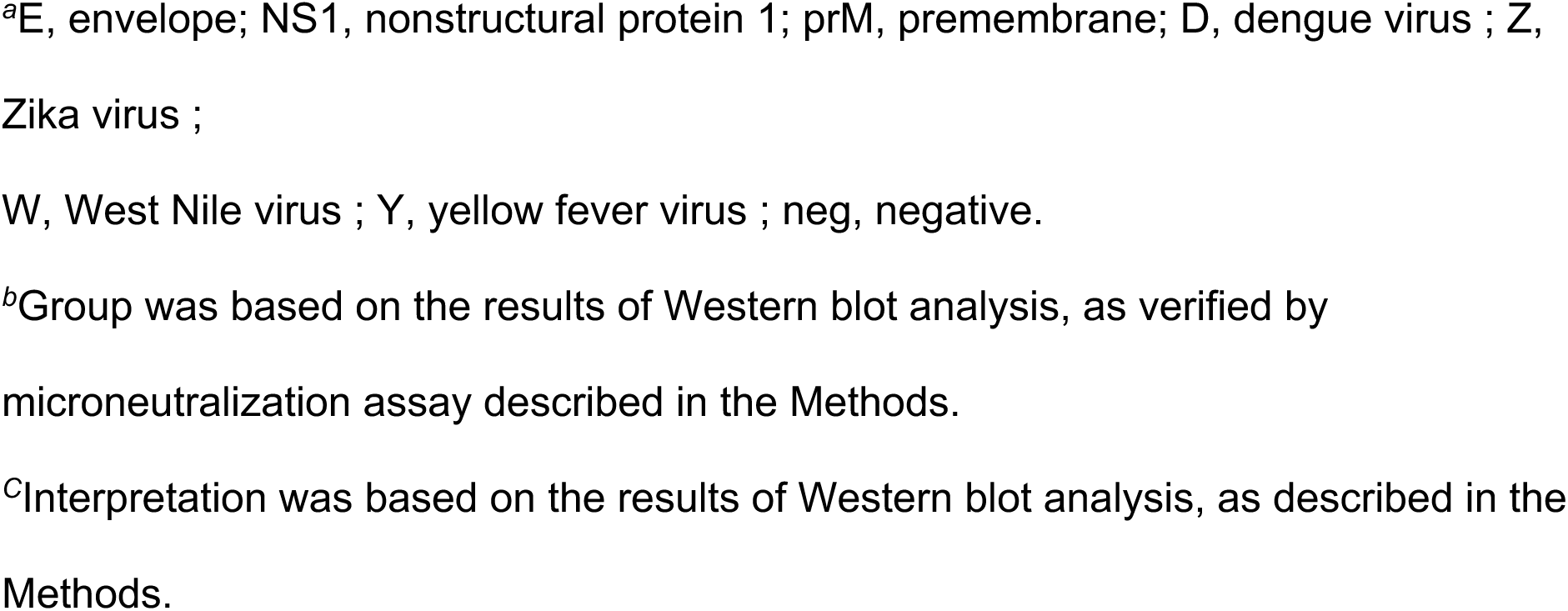
Summary of results of Western blot analysis.

**Supplementary Table S2.** Basic information of study participants compared to study population in Saúde, Brazil and seroprevalence to DENV, ZIKV and YFV among different age groups and gender.

| Groups<sup>a</sup> | Saude population<sup>b</sup><br>No. (%) | Study participants<br>No. (%) | Seroprevalence<sup>c</sup><br>No. of positive/samples tested (%) |  |  |
| --- | --- | --- | --- | --- | --- |
|  |  |  | DENV | ZIKV | YFV |
| total | 10,478 (100.0%) | 300 (100.0%) | 213/300<br>(71.0%) | 61/300<br>(20.3%) | 175/300<br>(58.3%) |
| Age group |  |  |  |  |  |
| ≤19 | 3,011 (28.7%) | 22 (7.3%) | 17/22<br>(77.3%) | 4/22<br>(18.2%) | 16/22<br>(72.7%) |
| 20–39 | 2,662 (25.4%) | 123 (41.0%) | 83/123<br>(67.5%) | 22/123<br>(17.9%) | 68/123<br>(55.3%) |
| 40–59 | 2,785 (26.6%) | 103 (34.3%) | 77/103<br>(74.8%) | 26/103<br>(25.2%) | 61/103<br>(59.2%) |
| 60–79 | 1,654 (15.8%) | 43 (14.3%) | 29/43<br>(67.4%) | 8/43<br>(18.6%) | 26/43<br>(60.5%) |
| ≥80 | 366 (3.5%) | 9 (3.0%) | 7/9<br>(77.8%) | 1/9<br>(11.1%) | 4/9<br>(44.4%) |
| Gender |  |  |  |  |  |
| male | 5,147 (49.1%) | 166 (55.3%) | 109/166<br>(65.7%) | 38/166<br>(22.9%) | 91/166<br>(54.8%) |
| female | 5,331 (50.9%) | 134 (44.7%) | 104/134<sup>d</sup><br>(77.6%) | 23/134<br>(17.2%) | 84/134<br>(62.7%) |

<sup>a</sup>Basic information includes total numbers, age groups and gender of study population in Saúde and study participants.
<sup>b</sup>Information based on Census data in 2022.
<sup>c</sup>Based on Western blot analysis (66).
<sup>d</sup>Comparing male and female, $P=0.02$ , Fisher's exact test.

## Supplemenatary Figures

**Supplementary Figure S1.**
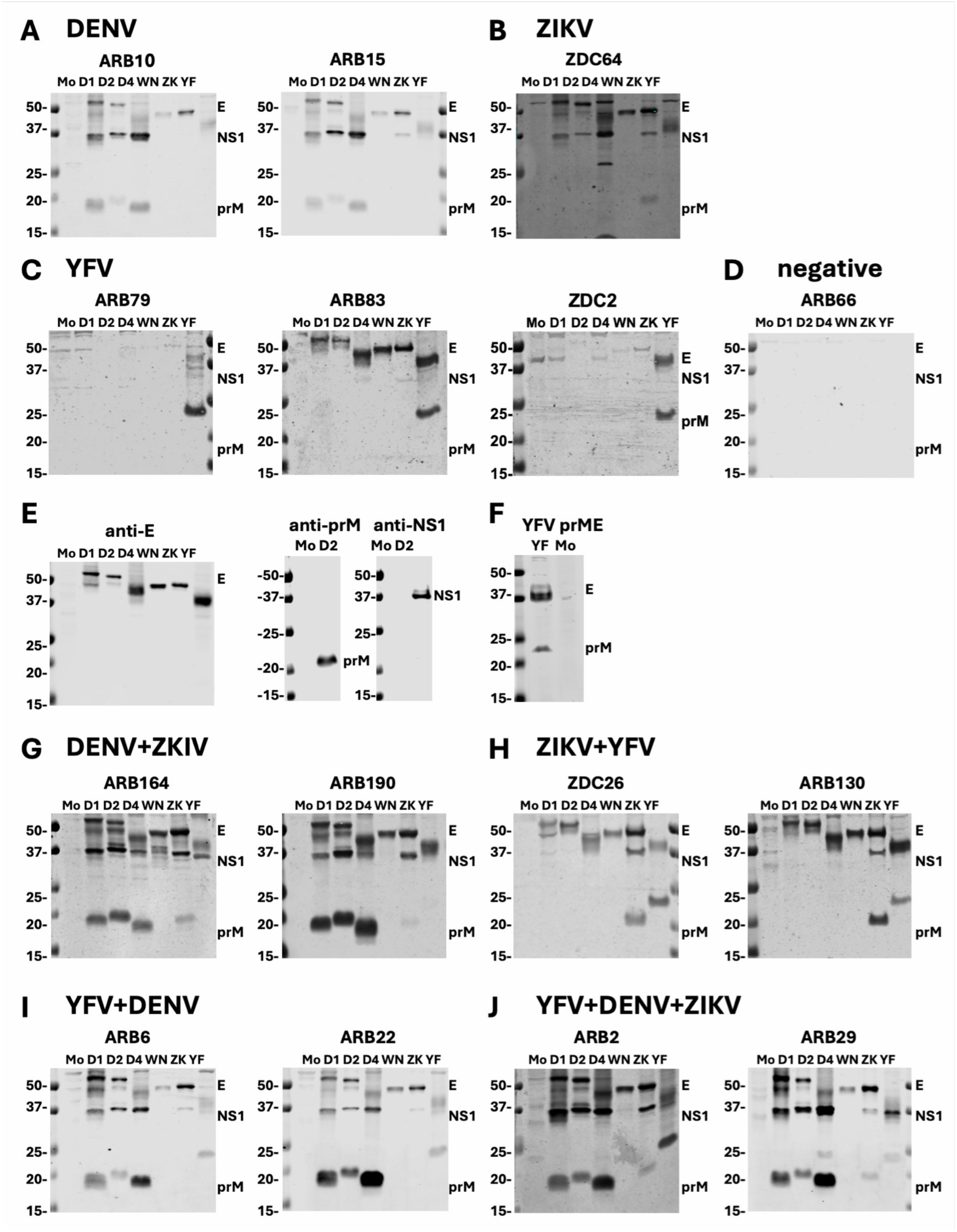
Previous orthoflavivirus infection/exposure in participants determined by Western blot analysis. Lysates derived from mock-, DENV1-, DENV2-, DENV4-, WNV-, ZIKV-, and YFV-17D-infected Vero cells were subjected to SDS-12% polyacrylamide gel electrophoresis under non-reducing condition and WB analysis probed with plasma samples or mouse monoclonal antibodies control (66). (A-D, G-J) Results of participants with previous DENV infection (A), ZIKV infection (B), YFV infection or vaccination (C), negative (D), DENV+ZIKV infections (G), ZIKV+YFV infections/vaccination (H), YFV+DENV infections/vaccination (I), and YFV+DENV+ZIKV infections/vaccination (J). (E) The blots were probed with anti-E (FL0232), anti-prM (70-12) or anti-NS1 (DB29-1) mouse monoclonal antibodies as described previously (10). (F) Lysates derived from 293T cells transfected with YFV-17D prME plasmid were subjected to Western blot analysis and probed with a YFV-17D vaccinee serum. The positions of E, NS1 and prM protein bands are indicated. The size of molecular weight markers is shown in kDa. Mo: mock, D1: DENV1, D2: DENV2, D4: DENV4, WN: WNV, ZK: ZIKV, and YF: YFV-17D.

**Supplementary Figure S2.**
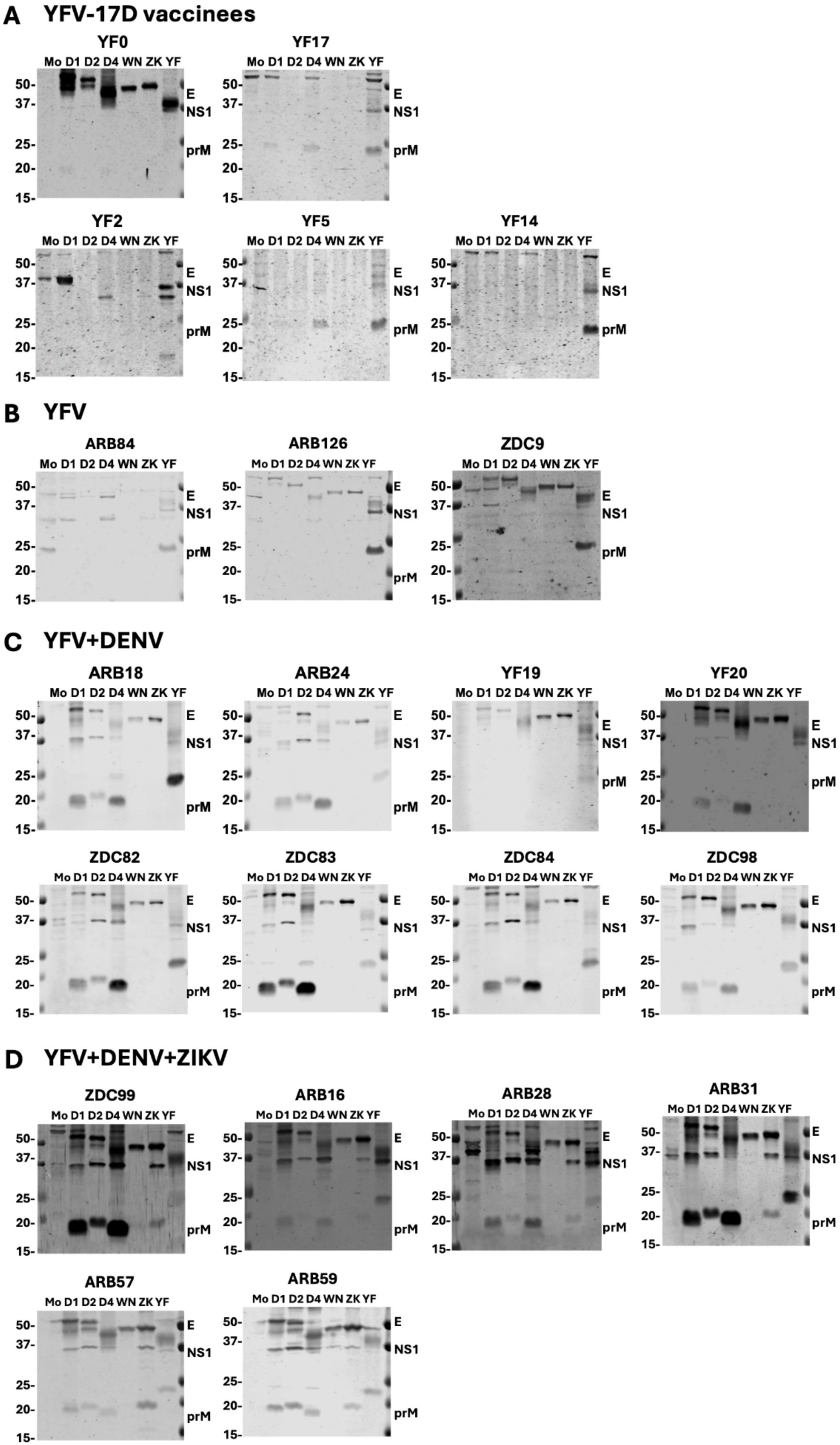
Western blot analysis of distinct YFV-exposed groups. Lysates derived from mock-, DENV1-, DENV2-, DENV4-, WNV-, ZIKV-, and YFV-17D-infected Vero cells were subjected to SDS-12% polyacrylamide gel electrophoresis under non-reducing condition and WB analysis probed with plasma samples. Results from additional participants with previous (A) YFV-17D vaccinees (YF0, YF17, YF2, YF5, and YF14), (B) YFV infection or vaccination (ARB84, ARB126, and ZDC9), (C) YFV+DENV infections/vaccination (ARB18, ARB24, YF19, YF20, ZDC82, ZDC83, ZDC84, and ZDC98), and (D) YFV+DENV+ZIKV infections/vaccination (ZDC99, ARB16, ARB28, ARB31, ARB57, and ARB59).

**Supplementary Figure S3.**
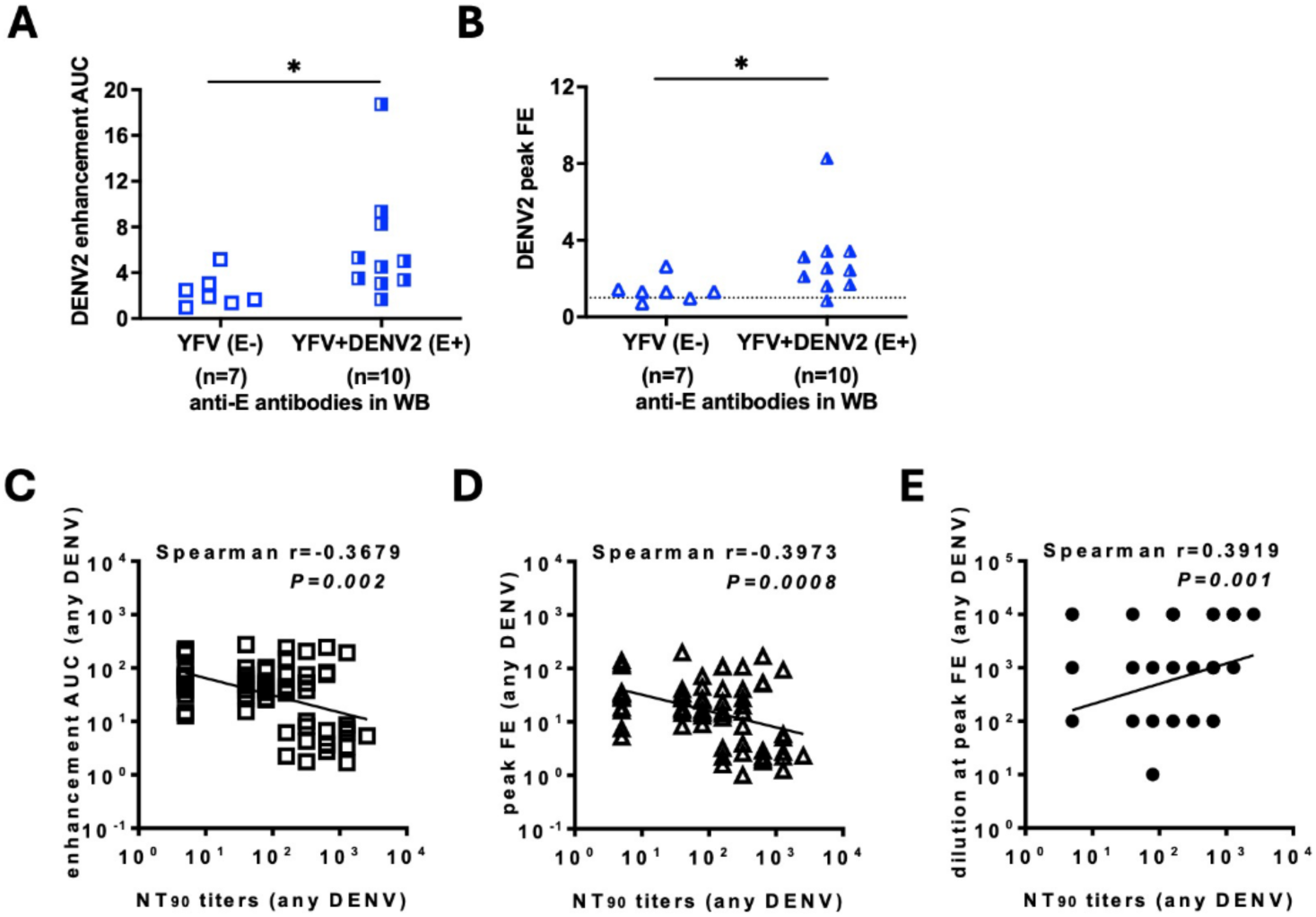
Relationship between ADE in vitro and binding antibody in YFV-only group and NT_90_ titers in YFV with multiple exposure groups. (A,B) Among YFV-only group, higher DENV2 enhancement was found in those with anti-E antibodies cross-reactive to DENV2 than those without based on area under the curve (AUC) (A) and peak fold enhancement (FE) (B) analysis. Dot lines indicate FE=1; Two-tailed Mann-Whitney U tests. (C,D,E) Among YFV with multiple exposure groups, the relationship between NT_90_ titers and enhancement AUC (C), peak FE (D), and plasma dilution at peak FE (E) considering all DENV serotypes together. Spearman correlation coefficient r.

**Supplementary Figure S4.**
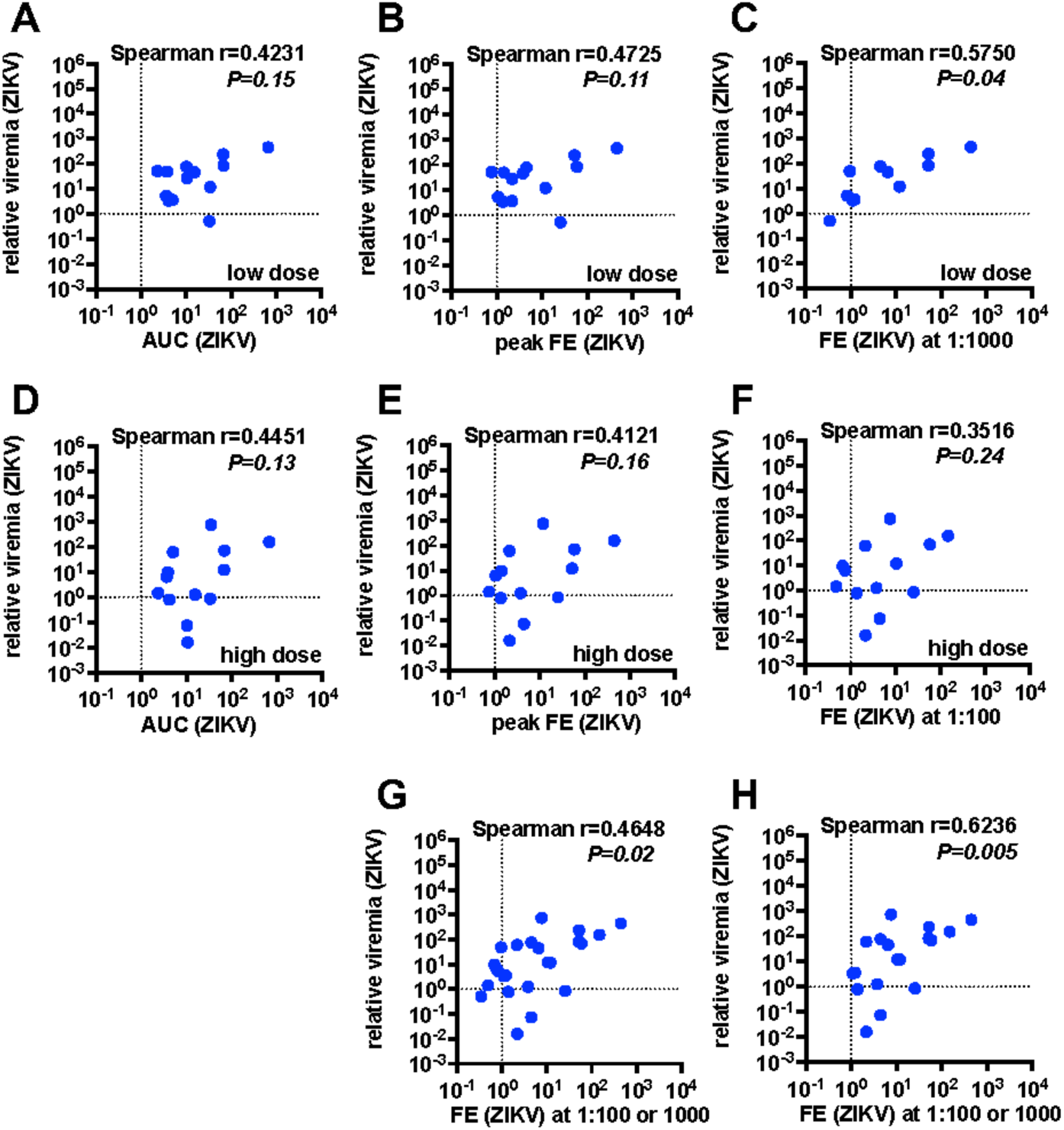
Relationship between ZIKV viremia in vivo and ADE activity in vitro for low dose and high dose passive transfers in ZIKV lethal challenge model. (A,B,C) The relationship between ZIKV relative viremia in vivo and ZIKV enhancement in vitro based on area under the curve (AUC) (A), peak fold enhancement (FE) (B), and FE at 1:1000 dilution (C) for the low dose transfer experiment. (D,E,F) The relationship between ZIKV relative viremia in vivo and ZIKV enhancement in vitro based on AUC (D), peak FE (E), and FE at 1:100 dilution (F) for the high dose transfer experiment. (G,H) The relationship between ZIKV relative viremia in vivo and ZIKV enhancement in vitro based on FE at 1:1000 or 1:100 dilution for both low and high dose transfer experiments together (G) and excluding those with FE<1 (H). ZIKV relative viremia=the ratio of RNA copies per µL in transfered mouse to the mean of RNA copies per µL in naive mice. Dot lines indicate FE=1 or relative viremia=1; Spearman correlation coefficient r; For low and high dose transfers, FE at 1:1000 and 1:100 were chosen, respectively, to mimic the dilution in vivo (2 µL and 100 µL/12.6 mL estimated blood volume of a 5-week-old *IFNAR1^-/-^* mouse).

**Supplementary Figure S5.**
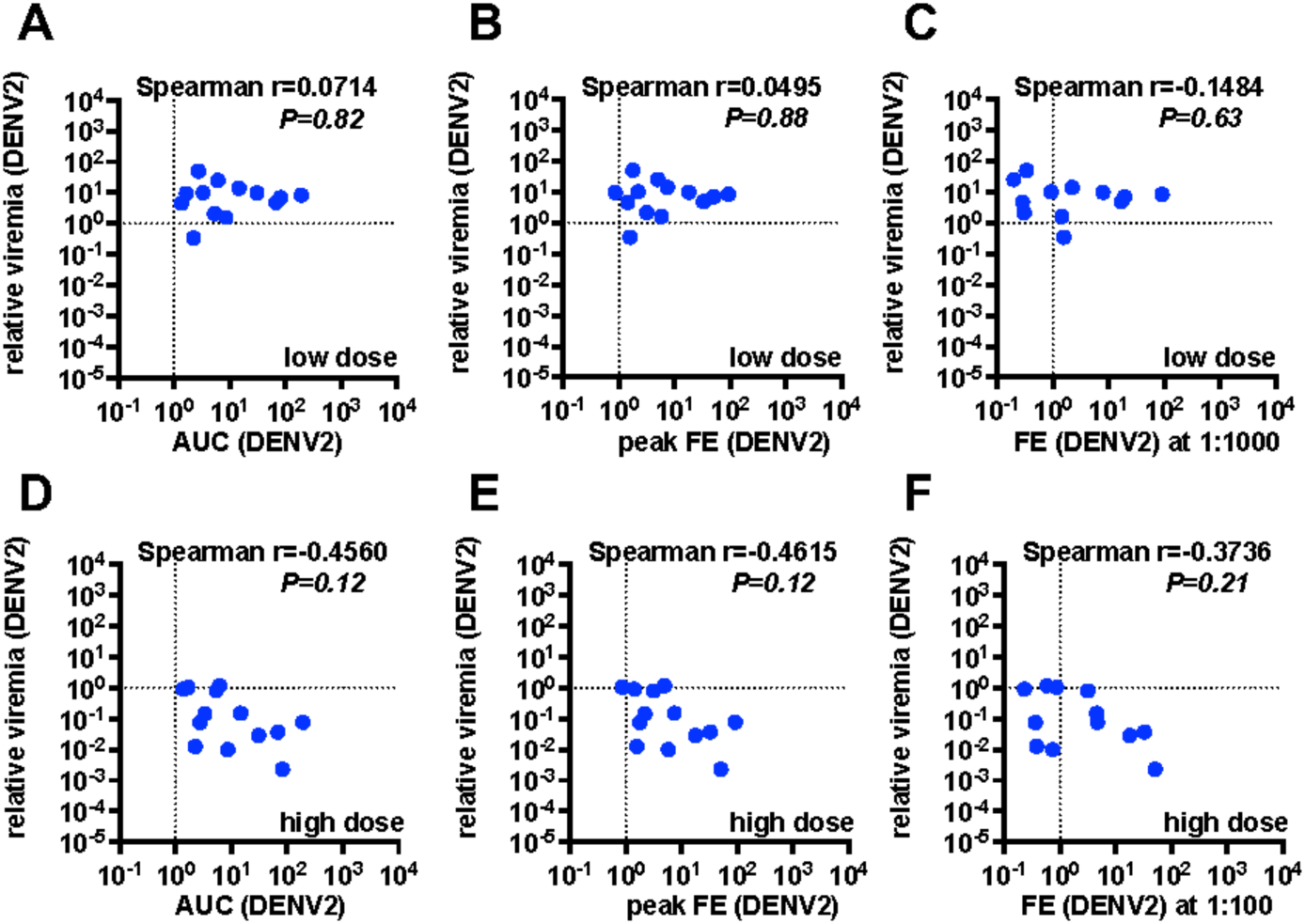
Relationship between DENV2 viremia in vivo and ADE activity in vitro for low dose and high dose passive transfers in DENV2 sublethal challenge model. (A,B,C) The relationship between DENV2 relative viremia in vivo and DENV2 enhancement in vitro based on area under the curve (AUC) (A), peak fold enhancement (FE) (B), and FE at 1:1000 dilution (C) for the low dose transfer experiment. (D,E,F) The relationship between ZIKV relative viremia in vivo and ZIKV enhancement in vitro based on AUC (D), peak FE (E), and FE at 1:100 dilution (F) for the high dose transfer experiment. DENV2 relative viremia=the ratio of RNA copies per µL in transferred mice to the mean of RNA copies per µL in naive mice. Dot lines indicate FE=1 or relative viremia=1; Spearman correlation coefficient r; For low and high dose transfers, FE at 1:1000 and :100 were chosen, respectively, to mimic the dilution in vivo (2 µL and 100 µL/12.6 mL estimated blood volume of a 5-week-old *IFNAR1^-/-^*mouse).

**Supplementary Figure S6.**
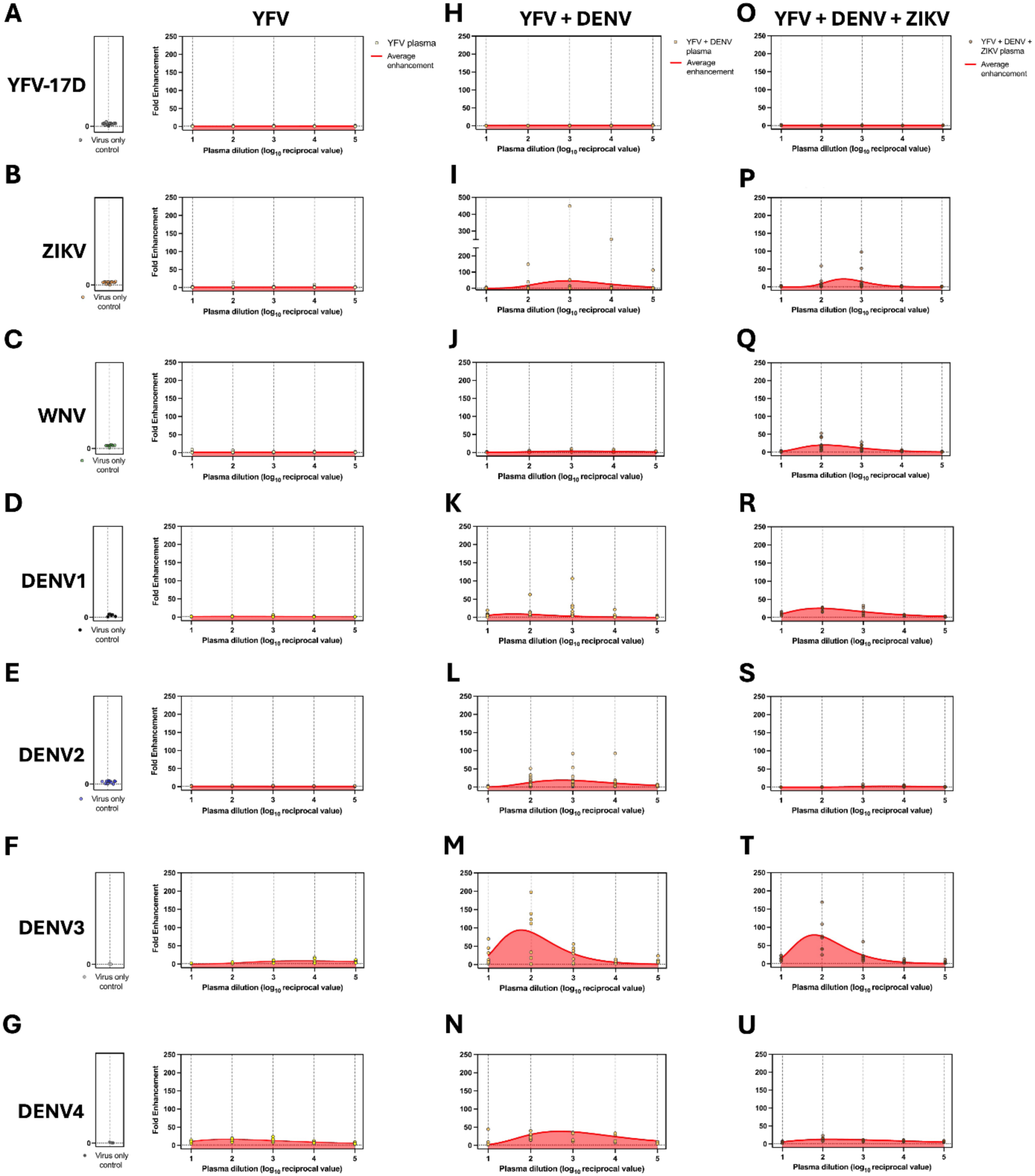
In vitro antibody-dependent enhancement of YFV-17D, ZIKV, WNV, and DENV1-4 across orthoflavivirus exposure groups. Serially diluted plasma from three groups, YFV-only (A-G), YFV+DENV (H-N), and YFV+DENV+ZIKV (O-U), were evaluated for Fcγ receptor-dependent antibody-dependent enhancement (ADE) using U937 cells. Plasma samples were incubated with virus prior to infection of U937 cells, and infection was quantified by intracellular staining for orthoflavivirus envelope protein. Enhancement of (A) YFV-17D, (B) ZIKV, (C) WNV, (D) DENV1, (E) DENV2, (F) DENV3, and (G) DENV4 is serially diluted plasma from individuals with a YFV-only profile. Enhancement of (H) YFV-17D, (I) ZIKV, (J) WNV, (K) DENV1, (L) DENV2, (M) DENV3, and (N) DENV4 is serially diluted plasma from individuals with a YFV+DENV profile. Enhancement of (O) YFV-17D, (P) ZIKV, (Q) WNV, (R) DENV1, (S) DENV2, (T) DENV3, and (U) DENV4 is serially diluted plasma from individuals with a YFV+DENV+ZIKV profile. Each symbol represents an individual plasma sample at the indicated dilution. The solid line denotes the group average FE across donors within each cohort. Virus-only controls consisted of challenge virus incubated with U937 cells in the absence of plasma and established baseline infectivity. FE for each plasma dilution was calculated as the ratio of infected cells relative to the corresponding virus-only control.

**Supplementary Figure S7.**
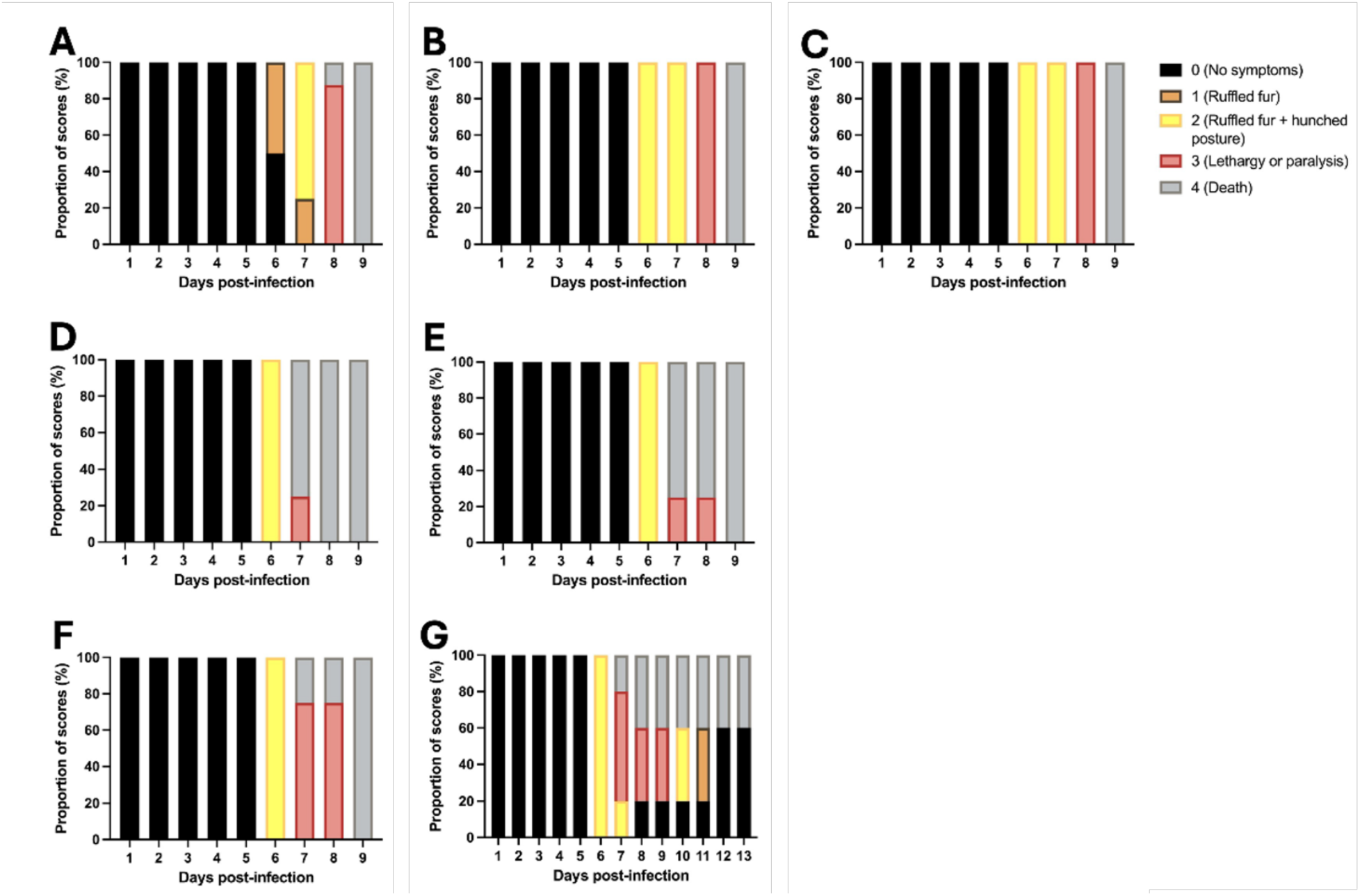
Clinical symptom progression following ZIKV challenge after passive transfer of human plasma. *IFNAR1^⁻/⁻^* mice were passively transferred with either 2 µL (low dose) or 100 µL (high dose) of human plasma 24 hours prior to ZIKV challenge and monitored daily for clinical signs. (A) Clinical scores of naïve plasma recipient control mice following 2 µL and 100 µL transfer. (B) Clinical scores of mice receiving 2 µL YFV-only plasma. (C) Clinical scores of mice receiving 100 µL YFV-only plasma. (D) Clinical scores of mice receiving 2 µL YFV+DENV plasma. (E) Clinical scores of mice receiving 100 µL YFV+DENV plasma. (F) Clinical scores of mice receiving 2 µL YFV+DENV+ZIKV plasma. (G) Clinical scores of mice receiving 100 µL YFV+DENV+ZIKV plasma. Clinical symptoms were assessed daily using a 5-point scale: 0, healthy; 1, ruffled fur or hunched posture; 2, ruffled fur and hunched posture; 3, ruffled fur, hunched posture, and lethargy or paralysis; 4, death. Mice were monitored through 9 days post infection (dpi) for naïve, YFV-only, and YFV+DENV groups, and through 9 dpi (2 µL) or 13 dpi (100 µL) for the YFV+DENV+ZIKV group, as indicated.

**Supplementary Figure S8.**
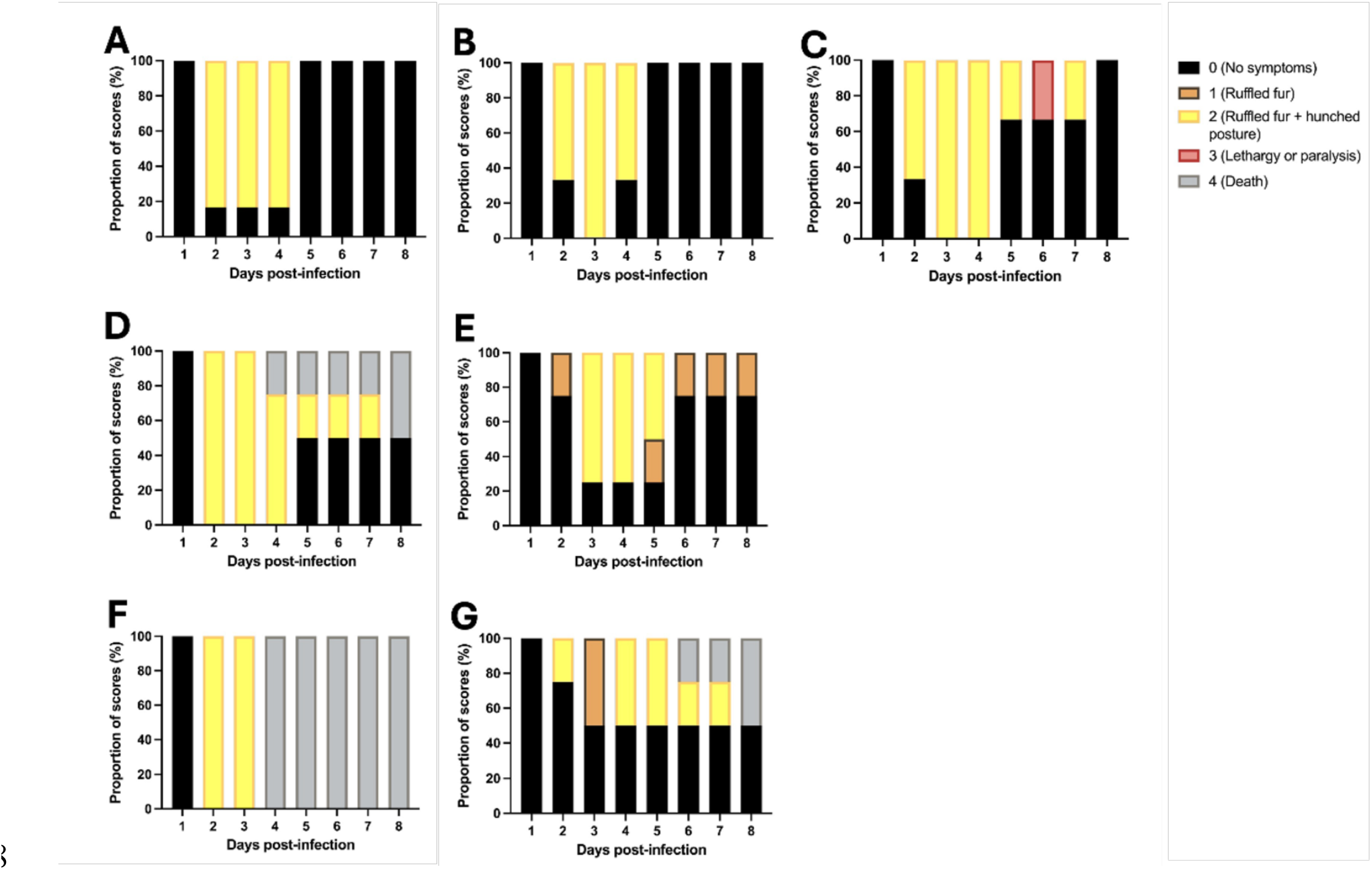
Clinical symptom progression following DENV2 challenge after passive transfer of human plasma. IFNAR1⁻/⁻ mice were passively transferred with either 2 µL (low dose) or 100 µL (high dose) of human plasma 24 hours prior to DENV2 challenge and monitored daily for clinical signs. (A) Clinical scores of naïve plasma recipient control mice following 2 µL and 100 µL transfer. (B) Clinical scores of mice receiving 2 µL YFV-only plasma. (C) Clinical scores of mice receiving 100 µL YFV-only plasma. (D) Clinical scores of mice receiving 2 µL YFV+DENV plasma. (E) Clinical scores of mice receiving 100 µL YFV+DENV plasma. (F) Clinical scores of mice receiving 2 µL YFV+DENV+ZIKV plasma. (G) Clinical scores of mice receiving 100 µL YFV+DENV+ZIKV plasma. Clinical symptoms were assessed daily using a 5-point scale: 0, healthy; 1, ruffled fur or hunched posture; 2, ruffled fur and hunched posture; 3, ruffled fur, hunched posture, and lethargy or paralysis; 4, death. Mice were monitored through 8 days post infection (dpi).

